# Epigenetic clocks of biological aging and risk of incident mild cognitive impairment and dementia: the Women’s Health Initiative Memory Study

**DOI:** 10.1101/2025.09.29.25336927

**Authors:** Steve Nguyen, Ake Lu, Steve Horvath, Mark A. Espeland, Stephen R. Rapp, Adam X. Maihofer, Caroline M. Nievergelt, Andrea Z. LaCroix, Linda K. McEvoy, Susan M. Resnick, Kenneth Beckman, Aladdin H. Shadyab

## Abstract

Aging is the strongest risk factor for dementia; however, few studies have examined the association of biological aging with incident dementia. We analyzed 6,069 cognitively unimpaired women (mean age=70.0 ± 3.8 years) in the Women’s Health Initiative Memory Study to examine the association of accelerated biological aging, measured with second and third-generation epigenetic clocks (AgeAccelPheno and AgeAccelGrim2, and DunedinPACE, respectively) with incident mild cognitive impairment (MCI) and probable dementia. Multivariable Cox proportional hazards models adjusted for age, race, ethnicity, smoking, hormone therapy regimen, physical activity, body mass index, and estimated white blood cell counts. For comparison, we also examined first-generation epigenetic clocks (AgeAccelHorvath; AgeAccelHannum). We evaluated effect modification by age, race/ethnicity, hormone therapy regimen, menopause type (natural vs surgical), and *APOE ε4* carriage. There were 1,307 incident MCI or probable dementia events over a median follow-up of 9.3 (25^th^ percentile=6.1, 75^th^ percentile=16.1) years. The adjusted HRs (95% CI; p-value) for incident MCI/probable dementia per one-standard deviation increment were 1.07 (1.01-1.15; p=0.03) for DunedinPACE, 1.11 (1.02-1.20; p=0.01) for AgeAccelGrim2, and 1.01 (0.95-1.07; p=0.74) for AgeAccelPheno. Only AgeAccelGrim2 was significant under the Bonferroni-corrected threshold for significance (p<0.02). Other epigenetic clocks were not associated with incident MCI/probable dementia. There was no effect modification in subgroup analyses (all P-interaction generally ≥ 0.05). In this cohort study of older women, accelerated biological aging measured by AgeAccelGrim2 was associated with higher risk of incident MCI/probable dementia. These findings provide evidence linking epigenetic biomarkers of biological aging with development of MCI and dementia independent of chronological age.

## INTRODUCTION

Aging is the strongest risk factor for Alzheimer’s disease (AD) and related dementias (“2024 Alzheimer’s Disease Facts and Figures,” 2024). Over two-thirds of AD cases are women. The estimated lifetime risk of AD among women aged 65 or older is 21.1% (nearly twice that of men at 11.6%), in part due to women living longer than men on average (Chene et al., 2015; Kochanek et al., n.d.). Thus, women represent an important population in which to further understanding of the heterogeneity of aging and dementia risk.

Epigenetic clocks are DNA methylation (DNAm)-based measures of biological aging that could inform on the heterogeneity of aging and dementia risk. Several studies have shown that epigenetic age acceleration, indicating faster biological aging relative to chronological age, is associated with higher risk of age-related outcomes including cardiovascular disease and mortality (Chen et al., 2016; Forrester et al., 2024). First-generation clocks including Horvath and Hannum DNAm age were trained on chronological age (Hannum et al., 2013; Horvath, 2013). Second-generation clocks including DNAm PhenoAge, DNAm GrimAge, and GrimAge2 were developed to predict phenotypic age and mortality, respectively (Levine et al., 2018; Lu et al., 2019, 2022, p. 2). Third-generation clocks, such as DunedinPACE, were developed to estimate the pace of biological aging using longitudinal data on biomarkers of organ system functioning (Belsky et al., 2022).

Yet, few large studies have examined the association of accelerated biological aging assessed with next-generation clocks, such as AgeAccelGrim2 and DunedinPACE, with incident mild cognitive impairment (MCI) and dementia. Prior studies had limited sample sizes, few MCI or dementia cases, less than 2 decades of follow-up, or examined few epigenetic clocks (Fransquet et al., 2021; Shadyab et al., 2022; Sugden et al., 2022; Thomas et al., 2024). Therefore, larger and more rigorous studies with longer follow-up periods are warranted.

We previously published preliminary findings suggesting potential associations of epigenetic age acceleration with cognitive outcomes among a subset of 578 women from the Women’s Health Initiative (WHI) Memory Study (WHIMS); however, we examined only four clocks and did not examine third-generation clocks (Shadyab et al., 2022). In this larger, well-powered study, we examined the associations of first-, second-and third-generation epigenetic clocks with incident MCI and probable dementia in 6,069 WHIMS women with up to 25 years of follow-up, performing the largest study on epigenetic clocks and incident MCI and probable dementia to date.

## METHODS

### Study population

The data supporting the present study’s findings are available on reasonable request from the WHI Program per publications and presentations policies (https://www.whi.org/doc/PP-policy.pdf).

WHIMS, an ancillary study of the WHI Hormone Trials, was designed to investigate the effects of hormone therapy (conjugated equine estrogens (CEE) alone vs. placebo or CEE plus medroxyprogesterone [E+P] acetate vs. placebo) on cognitive outcomes among 7,479 women aged 65-80 years recruited from 39 clinical sites across the U.S. who were cognitively unimpaired at randomization (1995-1998) (Shumaker et al., 1998). WHIMS administered annual in-person cognitive assessments during follow-up through 2007 for MCI and dementia (Shumaker et al., 1998). In 2008, WHIMS transitioned to annual telephone-based cognitive assessments in the WHIMS-Epidemiology of Cognitive Heath Outcomes (WHIMS-ECHO) study, with follow-up for MCI and probable dementia through 2021 (Espeland et al., 2017).

In the present study, we excluded 240 participants with only 1 WHIMS cognitive assessment, 519 who were ineligible for dbGaP, and 304 without available baseline DNA or buffy coat, resulting in 6,416 WHIMS women with DNAm data. After DNAm quality control (see Supplementary Methods), 347 women were excluded, resulting in 6,069 women who were free of MCI and probable dementia at baseline. The WHI Clinical Coordinating Center at Fred Hutchinson Cancer Research Center (Seattle, WA) approved all study protocols. This study was approved by the Institutional Review Board of University of California, San Diego. All women provided informed written consent. This study conforms to the Strengthening the Reporting of Observational Studies in Epidemiology (STROBE) guidelines.

### MCI and Probable Dementia Outcomes

Our primary outcome was the combined endpoint of MCI or probable dementia, whichever came first. We also examined MCI and probable dementia separately as secondary outcomes. Details on MCI and probable dementia adjudication in WHIMS are published elsewhere and briefly described below (Shumaker et al., 1998).

Participants completed the Modified Mini Mental State Examination (3MSE) in-person annually through 2007. Those scoring below age and education adjusted cutoffs completed a modified Consortium to Establish a Registry for Alzheimer’s Disease battery of neuropsychological and standardized tests (Shumaker et al., 1998). A physician at each study site with expertise in diagnosing dementia then classified women with MCI, probable dementia, or no cognitive impairment using Petersen’s criteria for MCI and Diagnostic and Statistical Manual of Mental Disorders, Fourth Edition criteria for probable dementia (American Psychiatric Association, 1994; Petersen et al., 2001). All participant data were sent to the WHIMS Clinical Coordinating Center for central adjudication by an expert panel consisting of a neurologist, geriatric psychiatrist, and geropsychologist (Shumaker et al., 1998).

WHIMS-ECHO used a battery of validated neurocognitive tests including the Telephone Interview for Cognitive Status-modified (TICS-m),(Espeland et al., 2017; Rapp et al., 2012) which measures global cognitive function similar to the 3MSE. For women who scored below age and education adjusted cutoffs on the TICS-m, an interview with the participant’s pre-identified proxy was conducted to obtain required information about her functional status. Otherwise, the protocol for diagnosing and centrally adjudicating MCI, probable dementia, or no cognitive impairment was the same as that of WHIMS.

The WHIMS-Supplemental Case Ascertainment Protocol (SCAP) was implemented to identify MCI and probable dementia cases in deceased or proxy-dependent participants (Gaussoin et al., 2012, 2019). The Dementia Questionnaire was administered to the participant-identified proxy respondent (Gaussoin et al., 2012, 2019). SCAP and prior WHIMS data were submitted to the central adjudication panel for final ascertainment of MCI/probable dementia, similar to WHIMS (Gaussoin et al., 2012, 2019).

### Epigenetic clocks

Details on DNAm measurement are provided in the Supplemental Methods. For our main analysis, we focused on 2 extensively studied second-generation clocks, AgeAccelPheno and AgeAccelGrim2, and a third-generation clock, DunedinPACE. We focused on these clocks because they have been shown in the literature to have stronger associations with age-related phenotypes compared with first-generation clocks (Belsky et al., 2022; Conole et al., 2025; Levine et al., 2018; Lu et al., 2022; Sugden et al., 2022). For comparison, we also examined 2 first-generation clocks, AgeAccelHorvath and AgeAccelHannum (Hannum et al., 2013; Horvath, 2013). In secondary analyses, we examined intrinsic and extrinsic epigenetic age acceleration (IEAA and EEAA, respectively), principal component (PC)-based versions of the first-and second-generation clocks (PCHorvath, PCHannum, PCPhenoAge, and PCGrimAge,), and the 10 DNAm-based AgeAccelGrim2 components individually (Higgins-Chen et al., 2022; Lu et al., 2022, p. 2). Details on calculation of the epigenetic clocks are provided in the Supplemental Methods.

For the epigenetic clocks, higher values indicate accelerated biological aging relative to chronological age. For DunedinPACE, values >1 indicate faster pace of aging (e.g., a value of 1.10 indicates a pace of aging 10% faster than the average), while values <1 indicate slower pace of aging (e.g., a value of 0.90 indicates a pace of aging 10% slower than the average) (Belsky et al., 2022).

### Covariates

Baseline questionnaires assessed age, race, ethnicity, education, smoking status, and total energy expenditure from recreational physical activity (metabolic equivalent hours/week). Height was measured with a stadiometer and weight with a balance beam scale to calculate body mass index (BMI; kg/m^2^). Menopause type was classified as natural or surgical (Shadyab et al., 2017). We defined *APOE ε4* carriage as presence of at least 1 *ε4* allele, which was determined among White women with available genome-wide genotyping data based on 2 single nucleotide polymorphisms: rs429358 and rs7412. Imputation was performed using the 1000 Genomics Project reference panel, and MaCH algorithms were implemented in Minimac (Howie et al., 2012). White blood cell (WBC) counts were estimated using IDOL (Salas et al., 2022).

### Statistical analysis

We calculated means and standard deviations for continuous variables or counts and proportions for categorical variables. We compared baseline characteristics across quartiles of the three clocks of primary interest in our study, including AgeAccelGrim2, DunedinPACE, and AgeAccelPheno separately, using F-tests for continuous variables or chi-square tests for categorical variables. We also compared characteristics between participants included in the analytic sample (n=6069) and WHIMS women who were excluded (n=1410). We calculated Pearson correlations between all epigenetic clocks. We estimated Kaplan-Meier curves across tertiles of each of the 5 main clocks we examined and capped follow-up at the 90^th^ percentile due to smaller risk sets from censoring and unstable survival estimates. Differences in survival across tertiles were assessed using log-rank tests.

Multivariable Cox proportional hazards regression models estimated hazard ratios (HR) and 95% confidence intervals (CI) for the associations of one-standard deviation (SD) increments in epigenetic clocks to facilitate comparisons of study results across clocks with different units of measurement, with the main endpoint of combined MCI/probable dementia and secondary outcomes of MCI and probable dementia separately. We calculated p-values for epigenetic clock associations using the Wald test for the regression coefficient in the models. We calculated follow-up time as days from baseline to the date of the cognitive assessment that triggered the first diagnosis of MCI or probable dementia (whichever came first for combined MCI/probable dementia), or the date of the last cognitive assessment, similar to prior WHIMS studies (Shumaker et al., 2003, 2004). For participants in WHIMS-SCAP, those with MCI or probable dementia had their follow-up time set to the midpoint of their last cognitive assessment and midpoint of the Dementia Questionnaire if they were alive, or date of death (Gaussoin et al., 2012, 2019). For those without MCI or probable dementia from WHIMS-SCAP, follow-up time was set to the maximum of the date of their last cognitive assessment or Dementia Questionnaire if they were alive, or date of death (Gaussoin et al., 2012, 2019). Schoenfeld residuals tested the proportional hazards assumption; no violations were observed.

Model 1 adjusted for age. Model 2 additionally included race and ethnicity as separate variables including all categories, as published studies have shown differences in epigenetic clocks, MCI, and dementia by race and ethnicity (Crimmins et al., 2021; Diaz-Venegas et al., 2016; Kornblith et al., 2022; Wright et al., 2021). Model 3 additionally included potential confounders informed by the published literature: education, hormone therapy trial arm, smoking, BMI, and physical activity. The fully adjusted model (model 4) additionally included estimated WBC counts. Models with IEAA, IEAA.Hannum, or EEAA did not contain WBC counts, since IEAA was derived to be independent of WBC counts, while EEAA incorporates them into its calculation. To account for missing covariate data, we applied multiple imputation by chained equations using the R *mice* package, specifying all study variables with 100 imputations and 5 iterations.

In sensitivity analyses, we repeated the main models excluding women who had MCI/probable dementia during the first 2 years of follow-up to mitigate the potential of reverse causation bias on the primary results. To evaluate the consistency of associations across cohort subgroups, we performed stratified analyses across age, hormone therapy trial arm, menopause type (natural vs surgical), race/ethnicity (Black, Hispanic or Latina, or White), and *APOE ε4* carriage. We statistically evaluated effect modification using cross-product interaction terms between epigenetic clocks and stratification variables.

We report nominal p-values for all tests. For completeness, we note that the Bonferroni-corrected threshold for the 3 main clocks of interest (AgeAccelPheno, AgeAccelGrim2, and DunedinPACE) and the primary outcome of MCI/probable dementia would be 0.0167 (0.05/3). We analyzed the data using R 4.4.1 from September 2024-September 2025.

## RESULTS

At baseline, women were 70.0 years on average, 0.2% were American Indian or Alaskan Native, 1.7% were Asian, 0.1% were Native Hawaiian or other Pacific Islander, 6.9% were Black, 89% were White, 1.1% reported more than one race, and 0.9% were unknown or not reported race (Table 1). Overall, 2.6% reported Hispanic or Latina ethnicity. Compared with women in the lowest quartile of AgeAccelGrim2, those in the highest quartile were less likely to be White, less likely to have higher educational attainment, more likely to smoke, had higher BMI, and reported less physical activity (Table 1). A generally similar pattern of differences across quartiles was noted for DunedinPACE and AgeAccelPheno (eTables 1 and 2). However, women in the highest quartile of AgeAccelPheno were more likely to be White compared to those in the lowest (eTable 2). Compared with women in the analytic sample, women who were excluded were older, were more likely to be Hispanic or Latina, were less likely to have high educational attainment, and were similar across race, smoking, BMI, physical activity, and *APOE ε4* carriage (eTable 3).

**Table 1.** Baseline sociodemographic, behavior, and health characteristics by quartiles of AgeAccelGrim2 in the Women’s Health Initiative Memory Study (n=6069)

| Characteristics | Total | AgeAccelGrim2 quartiles |  |  |  | p-value |
| --- | --- | --- | --- | --- | --- | --- |
|  |  | Q1 (n=1518):<br>[-14.4, -2.97] | Q2 (n=1517):<br>(-2.97, -0.56] | Q3 (n=1517):<br>(-0.56, 2.30] | Q4<br>(n=1517):<br>(2.30, 21.7) |  |
|  | No. (%) | No. (%) | No. (%) | No. (%) | No. (%) |  |
| Age, *mean (sd)* | 70.0 (3.8) | 69.8 (3.7) | 70.0 (3.8) | 70.5 (3.9) | 69.9 (3.9) | <0.001 |
| Highest education level, *n (%)* |  |  |  |  |  | <0.001 |
| Less than high school/GED | 421 (7) | 76 (5) | 93 (6.1) | 100 (6.6) | 152 (10.1) |  |
| High school/GED | 1306 (21.6) | 332 (22) | 324 (21.4) | 331 (21.9) | 319 (21.1) |  |
| Some college | 2414 (39.9) | 568 (37.6) | 593 (39.2) | 612 (40.5) | 641 (42.5) |  |
| College graduate or higher | 1906 (31.5) | 536 (35.4) | 504 (33.3) | 468 (31) | 398 (26.4) |  |
| Hormone trial arm |  |  |  |  |  | 0.003 |
| Estrogen alone intervention | 1152 (19) | 250 (16.5) | 301 (19.8) | 304 (20) | 297 (19.6) |  |
| Estrogen placebo | 1163 (19.2) | 253 (16.7) | 298 (19.6) | 289 (19.1) | 323 (21.3) |  |
| Estrogen plus progestin intervention | 1854 (30.5) | 489 (32.2) | 456 (30.1) | 456 (30.1) | 453 (29.9) |  |
| Estrogen plus progestin placebo | 1900 (31.3) | 526 (34.7) | 462 (30.5) | 468 (30.9) | 444 (29.3) |  |
| Race, *n (%)* |  |  |  |  |  | <0.001 |
| American Indian or Alaskan Native | 13 (0.2) | 4 (0.3) | 4 (0.3) | 1 (0.1) | 4 (0.3) |  |
| Asian | 106 (1.7) | 32 (2.1) | 29 (1.9) | 24 (1.6) | 21 (1.4) |  |
| Native Hawaiian or other Pacific Islander | 8 (0.1) | 2 (0.1) | 2 (0.1) | 2 (0.1) | 2 (0.1) |  |
| Black | 417 (6.9) | 54 (3.6) | 74 (4.9) | 115 (7.6) | 174 (11.5) |  |
| White | 5403 (89) | 1400 (92.2) | 1380 (91) | 1343 (88.5) | 1280 (84.4) |  |
| More than 1 race | 68 (1.1) | 14 (0.9) | 14 (0.9) | 20 (1.3) | 20 (1.3) |  |
| Unknown or not reported | 54 (0.9) | 12 (0.8) | 14 (0.9) | 12 (0.8) | 16 (1.1) |  |
| Ethnicity, *n (%)* |  |  |  |  |  | 0.15 |
| Not Hispanic or Latina | 5890 (97.4) | 1482 (98.1) | 1473 (97.2) | 1462 (96.6) | 1473 (97.5) |  |
| Hispanic or Latina | 160 (2.6) | 29 (1.9) | 42 (2.8) | 51 (3.4) | 38 (2.5) |  |
| Unknown or not reported | 19 (0.3) | 7 (0.5) | 2 (0.1) | 4 (0.3) | 6 (0.4) |  |
| **Health behaviors/status** |  |  |  |  |  |  |
| Smoking status, *n (%)* |  |  |  |  |  | <0.001 |
| Never smoked | 3187 (53.3) | 1094 (72.8) | 970 (64.7) | 808 (54.3) | 315 (21.2) |  |
| Past smoker | 2379 (39.8) | 405 (27) | 513 (34.2) | 642 (43.1) | 819 (55.1) |  |
| Current smoker | 412 (6.9) | 3 (0.2) | 17 (1.1) | 39 (2.6) | 353 (23.7) |  |
| Body mass index; kg/m<sup>2</sup>, *mean (SD)* | 28.6 (5.7) | 27.1 (5.2) | 28.5 (5.4) | 29.3 (5.8) | 29.3 (6.0) | <0.001 |
| Total energy expenditure from recreational physical activity; MET-hours/week, *mean (SD)* | 11.4 (13.2) | 13.4 (14.0) | 12.0 (14.1) | 10.4 (12.2) | 9.6, (12.2) | <0.001 |
| APOE ε4 carriage | 1306 (25.3) | 348 (25.7) | 307 (23.3) | 322 (25.2) | 329 (27.1) | 0.18 |

Abbreviations: MCI=mild cognitive impairment; IEAA=intrinsic epigenetic age acceleration;
EEAA=extrinsic epigenetic age acceleration
Intervals are expressed as (a, b], where $a < x \leq b$ ; the Q1 interval includes the minimum value.

### Associations of epigenetic clocks with cognitive outcomes

There were 1307 incident combined MCI/probable dementia events, including 799 incident MCI and 786 incident probable dementia events, over a median follow-up of 9.3 years (25^th^ and 75^th^ percentiles=6.1 and 16.1 years, respectively). Pearson correlations between epigenetic clocks ranged from 0.03 to 0.96 (eFigure 1). Kaplan-Meier curves for the epigenetic clocks and the cognitive outcomes are shown in eFigures 2-6.

In fully adjusted models, every one SD increment in DunedinPACE (corresponding to an 11% faster pace of aging compared with the average) was associated with higher risk of the combined endpoint of incident MCI/dementia (HR=1.07, 95% CI=1.01-1.15; p=0.03) and MCI (HR=1.11, 95% CI=1.02-1.21; p=0.01) but not probable dementia (HR=1.01, 95% CI=0.93-1.10; p=0.82). Every one SD increment in AgeAccelGrim2 (4.29 years) was associated with higher risk of incident MCI/probable dementia (HR=1.11, 95% CI=1.02-1.20; p=0.01) but not probable dementia (HR=1.11, 95% CI=1.00-1.23; p=0.05) or MCI (HR=1.10, 95% CI=1.00-1.22; p=0.06; Figure 1). Each one-standard deviation increment in AgeAccelPheno was not associated with risk of MCI/probable dementia (HR=1.01, 95% CI=0.95-1.07; p=0.74), MCI (HR=0.99, 95% CI=0.92-1.07; p=0.89), or probable dementia (HR=1.02, 95% CI=1.02-0.94-1.10; p=0.62; Figure 1). Only the association between AgeAccelGrim2 and the primary outcome of incident MCI/probable dementia was considered significant after applying the Bonferroni-corrected threshold of 0.0167; the association for DunedinPACE was not significant at this threshold. Results from minimally and progressively adjusted models are shown in eTable 4.

**Figure 1.**
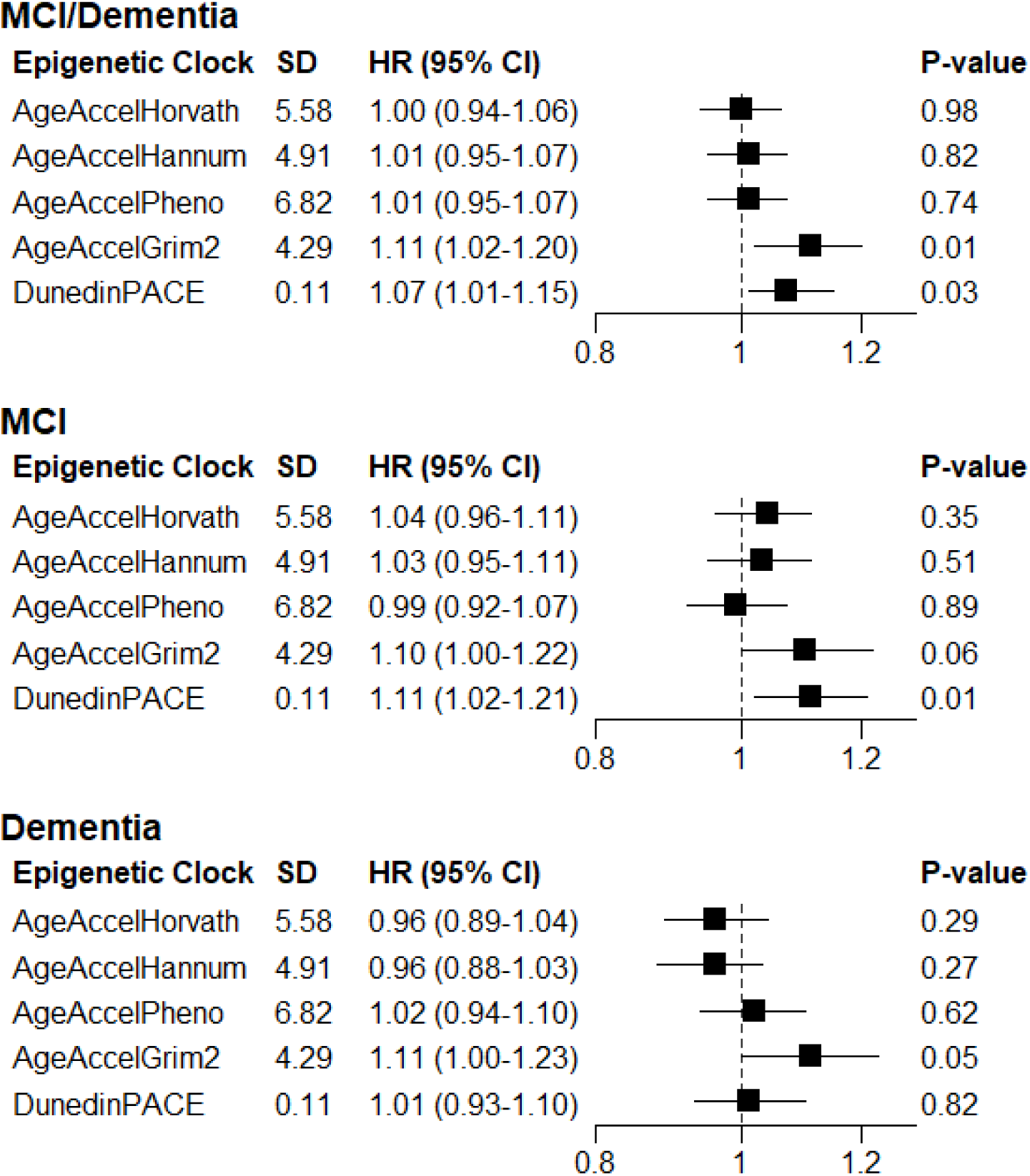
Associations of one-standard deviation increases in epigenetic clocks with incident mild cognitive impairment (MCI)/probable dementia, MCI, and probable dementia in the Women’s Health Initiative Memory Study (WHIMS; n=6069) 1995-2021. Abbreviations: SD=standard deviation; MCI=mild cognitive impairment; HR=hazard ratio; CI=confidence interval There were 1307 incident MCI/dementia events, 799 incident MCI events, and 786 incident probable dementia events in an analytic sample of 6069 for analyses with AgeAccelHorvath, AgeAccelHannum, IEAA, EEAA, IEAA.Hannum, AgeAccelPheno, and AgeAccelGrim2. There were 1301 incident MCI/dementia events, 797 incident MCI events, and 782 incident probable dementia events in an analytic sample of 6026 for analyses with DunedinPACE. The sum of separate incident MCI and probable dementia events is higher than the 1307 incident MCI/probable dementia events because participants with incident MCI later had incident probable dementia. Models adjusted for age, education, smoking, race, ethnicity, hormone therapy trial arm, total energy expenditure from recreational physical activity, body mass index, and white blood cell counts (CD8T, CD4T, natural killer cells, B cells, monocytes, and neutrophils). Models with IEAA, EEAA, and IEAA.Hannum did not include white blood cell counts. Missing covariate data were imputed using multiple imputation by chained equations with the R mice package.

In secondary analyses, every one SD increment in the PC-based version of AgeAccelGrim, PCGrimAgeResid (3.34 years), was associated with higher risk of MCI/probable dementia (HR=1.11, 95% CI=1.02-1.21; p=0.01) and MCI (HR=1.13, 95% CI=1.02-1.26; p=0.02) but not probable dementia in the fully adjusted models (eTable 4, eFigure 7). None of the other epigenetic clocks was associated with the cognitive outcomes in the fully adjusted models (eTable 4, eFigure 7).

In analyses of AgeAccelGrim2 components, higher DNAm PAI-1 was associated with higher risk of incident MCI/probable dementia (HR=1.10, 95% CI=1.04-1.17; p=0.002) and MCI (HR=1.10, 95% CI=1.02-1.19; p=0.01) but not probable dementia (eFigure 7). Higher DNAm log-HbA1C was associated with higher risk of MCI/probable dementia (HR=1.07, 95% CI=1.00-1.13; p=0.04) but not with MCI or probable dementia separately (eFigure 7). Higher DNAm PACKYRS was associated with higher risk of probable dementia (HR=1.13, 95% CI=1.01-1.26, p=0.03) but not MCI or MCI/probable dementia. The other AgeAccelGrim2 components were not associated with these outcomes (eFigure 7, eTable 4). Results from sensitivity analysis that excluded data from 79 women who developed MCI/probable dementia during the first 2 years of follow-up were consistent in direction and magnitude with results from the main analyses (eFigure 8).

In stratified analyses, associations of DunedinPACE and AgeAccelGrim2 with MCI/probable dementia generally did not vary by age, race, menopause type, hormone therapy regimen, or APOE ε4 carriage (Figures 2 and 3, and eTable 5).

**Figure 2.**
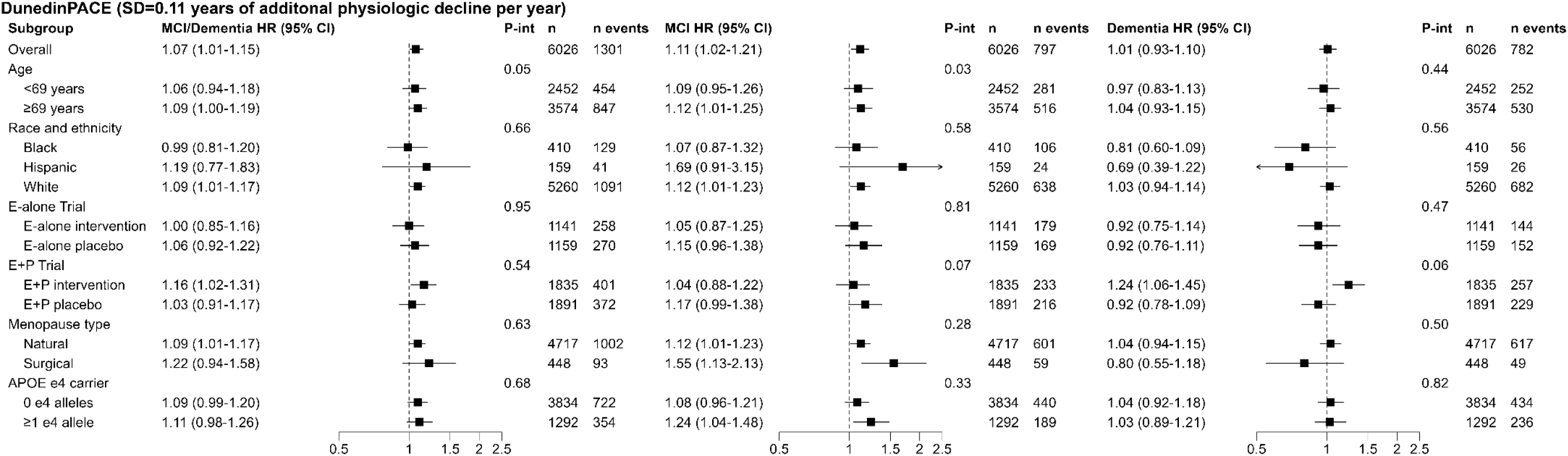
Associations of DunedinPACE with incident mild cognitive impairment (MCI)/probable dementia, MCI, and probable dementia stratified by age, race/ethnicity, hormone therapy regimen, menopause type, and *APOE ε4* carriage in the Women’s Health Initiative Memory Study (n=6026), 1995-2021. Abbreviations: HR = hazard ratio, CI = confidence interval, P-int=P-interaction, BMI = body mass index, E=conjugated equine estrogens alone, E+P=conjugated equine estrogens plus medroxyprogesterone acetate. Models adjusted for age, race/ethnicity, education, hormone therapy trial arm, total energy expenditure from recreational physical activity, BMI, and white blood cell counts (CD8T, CD4T, natural killer cells, B cells, monocytes, and neutrophils). Models across strata of race and ethnicity, hormone therapy trial arm, menopause type, and APOE ε4 carriage did not contain the stratification variable.

**Figure 3.**
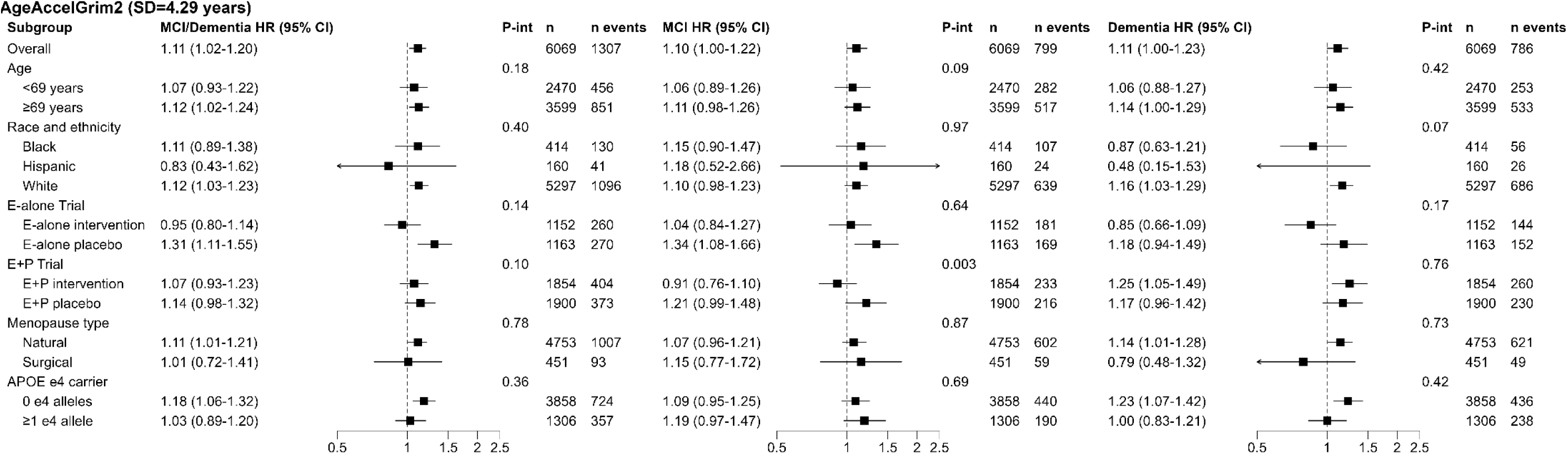
Associations of AgeAccelGrim2 with incident mild cognitive impairment (MCI)/probable dementia, MCI, and probable dementia stratified by age, race/ethnicity, hormone therapy regimen, menopause type, and *APOE ε4* carriage in the Women’s Health Initiative Memory Study (n=6,069), 1995-2021. Abbreviations: HR = hazard ratio, CI = confidence interval, P-int=P-interaction, BMI = body mass index, E=conjugated equine estrogens alone, E+P=conjugated equine estrogens plus medroxyprogesterone acetate. Models adjusted for age, race/ethnicity, education, hormone therapy trial arm, total energy expenditure from recreational physical activity, BMI, and white blood cell counts (CD8T, CD4T, natural killer cells, B cells, monocytes, and neutrophils). Models across strata of race and ethnicity, hormone therapy trial arm, menopause type, and APOE ε4 carriage did not contain the stratification variable.

## DISCUSSION

In a large cohort of older women, we performed the largest prospective study of epigenetic clocks and incident MCI and probable dementia to date. Our study showed that accelerated biological aging as measured by second-and third-generation clocks developed to predict mortality (AgeAccelgrim2) or the pace of aging (DunedinPACE) was associated with higher risk of incident MCI/probable dementia up to 26 years later independent of chronological age, sociodemographic characteristics, lifestyle behaviors, and health factors. After adjustment for multiple comparisons, only AgeAccelGrim2 remained significantly associated with incident MCI/probable dementia. We performed subgroup analyses by age, race/ethnicity, and *APOE* ε4 carriage, as well as menopause type and randomization to hormone therapy, which has not been previously evaluated, and found that associations generally did not differ across cohort subgroups. Other epigenetic clocks were not associated with these cognitive outcomes.

The varying associations of epigenetic clocks with cognitive outcomes observed in the present study could be attributable to the divergent methods used to develop each clock and the corresponding features of aging they may capture. DunedinPACE was developed from longitudinal data using 19 biomarkers indicative of organ system function, including the cardiovascular system and cardiometabolic health (Belsky et al., 2022). AgeAccelGrim2 was developed as a mortality estimator, incorporating DNA methylation associated with smoking, chronological age, and plasma proteins associated with mortality (Lu et al., 2022). In contrast, first generation epigenetic clocks were developed to predict chronological age (Bo et al., 2025; Horvath & Raj, 2018). Importantly, neither AgeAccelGrim2 nor DunedinPACE was developed to capture cognitive function or dementia risk, yet both showed associations with MCI/probable dementia (Belsky et al., 2022; Conole et al., 2025; Lu et al., 2022, p. 2). However, because DunedinPACE did not survive adjustment for multiple comparisons, replication in other studies is warranted. Overall, our findings suggest that AgeAccelGrim2 may capture biological processes that contribute to the development of MCI and dementia.

Few large studies have examined associations of epigenetic clocks with incident MCI/probable dementia. In the WHIMS cohort, we previously found that higher DunedinPACE was associated with faster cognitive decline as measured by the TICS-m test (Nguyen et al., 2024). In the Framingham Heart Study, a 0.12-unit increment in DunedinPACE was associated with 27% higher dementia risk, faster cognitive decline, and worse brain structural integrity (n=2,264, mean age=65, 54% women, 151 dementia events, 14-year follow-up), consistent in direction with our study findings (Savin et al., 2024; Sugden et al., 2022; Whitman et al., 2024). In the ASPirin in Reducing Events in the Elderly study (ASPREE), a 4-year increment in AgeAccelGrim2 was associated with 2-fold higher incident dementia risk among older men but not women (n=560, mean age∼74 years, ∼51% women, 87 dementia events, including 50 events among women, mean follow-up of 7 years) (Phyo et al., 2024). However, DunedinPACE was not associated with dementia risk in men or women (Phyo et al., 2024). Potential reasons for differences in findings across these studies and ours could be attributable to the larger study population of older women and longer follow-up (up to 26 years) in WHIMS and differences in ascertainment of cognitive outcomes.

Few studies have examined associations of AgeAccelGrim2 or its components with MCI or dementia, as this clock was more recently developed (Lu et al., 2022, p. 2). Among AgeAccelGrim2 components, only DNAm PAI-1 and DNAm log-CRP were associated with MCI/probable dementia, while DNAm PACKYRS was associated with probable dementia. PAI-1 was found to be higher in postmortem brains with AD than in control brains in a prior study (Barker et al., 2012). A meta-analysis of 13 studies found that higher CRP was associated with higher risk of AD (Darweesh et al., 2018). Smoking is an established risk factor for dementia (Livingston et al., 2024).

Study strengths include the large population of older Black, Hispanic/Latina, and White women, annual cognitive testing, and rigorously adjudicated MCI and probable dementia. The long follow-up period and corresponding large number of MCI and probable dementia events in WHIMS enhanced statistical power. We examined first-, second-, and third-generation epigenetic clocks. The WHI collected extensive sociodemographic, behavioral, and health information, enabling comprehensive adjustment for potential confounding.

### Limitations

Our study has several limitations. The present study was carried out in a public health randomized trial of women, which could limit the generalizability of results. However, the WHI Hormone Therapy Trials’ exclusion criteria were designed to be minimal in order for the study results to be maximally generalizable to women aged 50-79 (Hays et al., 2003). In WHIMS, cognitive assessments changed from the in-person 3MSE to the telephone-administered TICS-m after 2007. However, we previously showed that telephone-based cognitive assessments were valid and reliable and that telephone-administered and face-to-face cognitive tests yielded equivalent scores (Rapp et al., 2012). As cognitive assessments were annual, the exact timing of MCI and probable dementia events is unknown, a limitation shared by all dementia studies. However, as few participants missed cognitive assessments, we set the date of cognitive assessment as the event date as an approximation, consistent with prior studies (Dunk et al., 2024; Liu et al., 2022; Sachs et al., 2023). Missing outcome information and informative censoring could occur if participants did not attend cognitive assessments. To reduce this possibility, the WHIMS SCAP was implemented to reduce undercounting of probable dementia cases (Gaussoin et al., 2012, 2019). AD and vascular dementia were not classified in WHIMS after 2007, precluding examination of dementia subtypes. However, AD accounts for approximately 60-80% of dementia cases, and most individuals with AD have mixed pathology (“2024 Alzheimer’s Disease Facts and Figures,” 2024; Kapasi et al., 2017). Although the WHIMS women who were excluded in analyses were largely similar to those in the analytic sample, there were slight differences in age, education, hormone therapy trial regimen, and ethnicity; however, findings were independent of these factors. Future studies inclusive of men and individuals from underrepresented groups are needed to evaluate reproducibility of study findings. Given the observational design, we cannot establish causality. Because we examined multiple epigenetic clocks and not all associations survived correction for multiple comparisons, results should be interpreted with caution.

## Conclusions

In this cohort study of older women, we found that accelerated biological aging as measured by DunedinPACE and AgeAccelGrim2 was associated with higher risk of incident MCI/probable dementia, with AgeAccelGrim2 remaining significant after adjustment for multiple comparisons. The modest magnitude of observed associations suggests the possibility that stronger epigenetic biomarkers of cognitive aging will be discovered through training clocks specifically on cognitive changes and incident MCI and dementia. Overall, these findings provide evidence linking biological aging processes with mild cognitive impairment and dementia in older women.

## ACKNOWLEDGEMENTS

We thank the WHI participants, staff, and investigators. The short list of WHI investigators can be found at: https://www-whi-org.s3.us-west-2.amazonaws.com/wp-content/uploads/WHI-Investigator-Short-List.pdf. The full list of WHI Investigators can be found at the following site: http://www.whi.org/researchers/Documents%2520%2520Write%2520a%2520Paper/WHI%2520Investigator%2520Long%2520List.pdf

## AUTHORS’ CONTRIBUTIONS

S.N. contributed to the conceptualization and design of the study, conducted data analysis and data interpretation, and led manuscript preparation and manuscript revisions. A.L. and S.H. contributed to data analysis and manuscript preparation and manuscript review. M.A.E. contributed expertise for data analysis, data interpretation, and contributed to manuscript review and manuscript revision. S.R.R. provided expertise in design of the study, contributed to data interpretation, and contributed expertise for manuscript preparation. A.X.M. and C.M.N. contributed to data processing, data analysis, manuscript preparation, and manuscript review. A.Z.L. contributed to the conceptualization and design of the study, data analysis, data interpretation, manuscript writing, and manuscript revisions. L.K.M. contributed to the conceptualization and design of the study, data interpretation, provided expertise for manuscript preparation and revisions. S.M.R. contributed to data analysis, data interpretation, and manuscript writing. K.B. contributed to data generation and provided feedback for manuscript preparation. A.H.S. secured funding, led the conceptualization and design of the study, provided feedback and guidance on data analysis and data interpretation, and contributed and provided guidance for manuscript preparation and manuscript revisions.

## DATA AVAILABILITY

The data supporting the present study’s findings are available upon reasonable request and review of proposals submitted through the Women’s Health Initiative (WHI) study website at https://www.whi.org/propose-a-paper. Access is granted to investigators from recognized research institutions.

### Ethics Statement

This study was approved by the Institutional Review Board of University of California, San Diego. All women provided informed written consent.

