## supplemental_materials for "Epigenetic clocks of biological aging and risk of incident mild cognitive impairment and dementia: the Women’s Health Initiative Memory Study"

**Supplemental Material**

**eFigure 1. Pearson correlations between baseline epigenetic clocks in the Women’s Health Initiative Memory Study (n=6026).**

**eFigure 2.** **Kaplan Meier curves for mild cognitive impairment (MCI)/probable dementia, MCI, and probable dementia across tertiles of AgeAccelHorvath in the Women’s Health Initiative Memory Study (WHIMS; n=6069) 1995-2021.**

**eFigure 3. Kaplan Meier curves for mild cognitive impairment (MCI)/probable dementia, MCI, and probable dementia across tertiles of AgeAccelHannum in the Women’s Health Initiative Memory Study (WHIMS; n=6069) 1995-2021.**

**eFigure 4.** **Kaplan Meier curves for mild cognitive impairment (MCI)/probable dementia, MCI, and probable dementia across tertiles of AgeAccelPheno in the Women’s Health Initiative Memory Study (WHIMS; n=6069) 1995-2021.**

**eFigure 5. Kaplan Meier curves for mild cognitive impairment (MCI)/probable dementia, MCI, and probable dementia across tertiles of AgeAccelGrim2 in the Women’s Health Initiative Memory Study (WHIMS; n=6069) 1995-2021.**

**eFigure 6. Kaplan Meier curves for mild cognitive impairment (MCI)/probable dementia, MCI, and probable dementia across tertiles of DunedinPACE in the Women’s Health Initiative Memory Study (WHIMS; n=6069) 1995-2021.**

**eFigure 7.** **Associations of epigenetic clocks and AgeAccelGrim2 components with incident mild cognitive impairment (MCI)/probable dementia, MCI, and probable dementia in the Women’s Health Initiative Memory Study (WHIMS; n=6069) 1995-2021.**

**eFigure 8. Associations of epigenetic clocks and AgeAccelGrim2 components with incident mild cognitive impairment (MCI)/probable dementia, MCI, and probable dementia in the Women’s Health Initiative Memory Study after excluding data from 79 women who had incident MCI/probable dementia during the first 2 years of follow-up (WHIMS; n=5990) 1995-2021.**

**eFigure 9. Discriminatory accuracy of epigenetic clocks for incident mild cognitive impairment/probable dementia, (a) alone and (b) with the addition of age, race, and ethnicity.**

**eTable 1. Baseline sociodemographic, behavior, and health characteristics by quartiles of DunedinPACE in the Women’s Health Initiative Memory Study (n=6026)**

**eTable 2. Baseline sociodemographic, behavior, and health characteristics by quartiles of AgeAccelPheno in the Women’s Health Initiative Memory Study (n=6069)**

**eTable 3. Baseline sociodemographic, behavioral, and health characteristics of 6069 Women’s Health Initiative Memory Study women included in the analytic sample versus those excluded (n=1410)**

**eTable 4. Associations of epigenetic clocks and AgeAccelGrim2 components with incident mild cognitive impairment (MCI)/probable dementia, MCI, and probable dementia in the Women’s Health Initiative Memory Study (WHIMS; n=6069) 1995-2021.**

**eTable 5. Associations of epigenetic clocks with incident mild cognitive impairment (MCI)/probable dementia, MCI, and probable dementia stratified by age, race/ethnicity, hormone therapy regimen, menopause type, and APOE ε4 carriage in the Women’s Health Initiative Memory Study (n=6,069) 1995-2021.**

**Supplemental Methods**

**eFigure 1. Pearson correlations between baseline epigenetic clocks in the Women’s Health Initiative Memory Study (n=6026).**

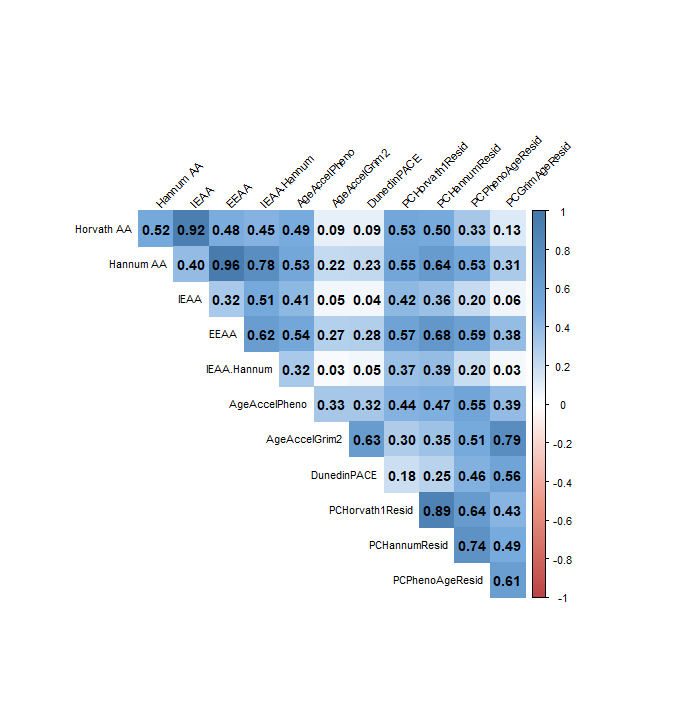
Abbreviations: AA = age acceleration, IEAA = intrinsic epigenetic age acceleration, EEAA = extrinsic epigenetic age acceleration
Horvath AA, Hannum AA, IEAA, EEAA, IEAA.Hannum, and AgeAccelGrim2 were calculated using the Horvath and Clock Foundation DNA Metyhlation age calculator available at <https://dnamage.clockfoundation.org/>. DunedinPACE was calculated using R code available at <https://github.com/danbelsky/DunedinPACE>. PCHorvath1Resid, PCHannumResid, PCPhenoAgeResid, and PCGrimAgeResid were calculated using R code available at <https://github.com/MorganLevineLab/PC-Clocks>.

**eFigure 2. Kaplan Meier curves for mild cognitive impairment (MCI)/probable dementia, MCI, and probable dementia across tertiles of AgeAccelHorvath in the Women’s Health Initiative Memory Study (WHIMS; n=6069) 1995-2021.**

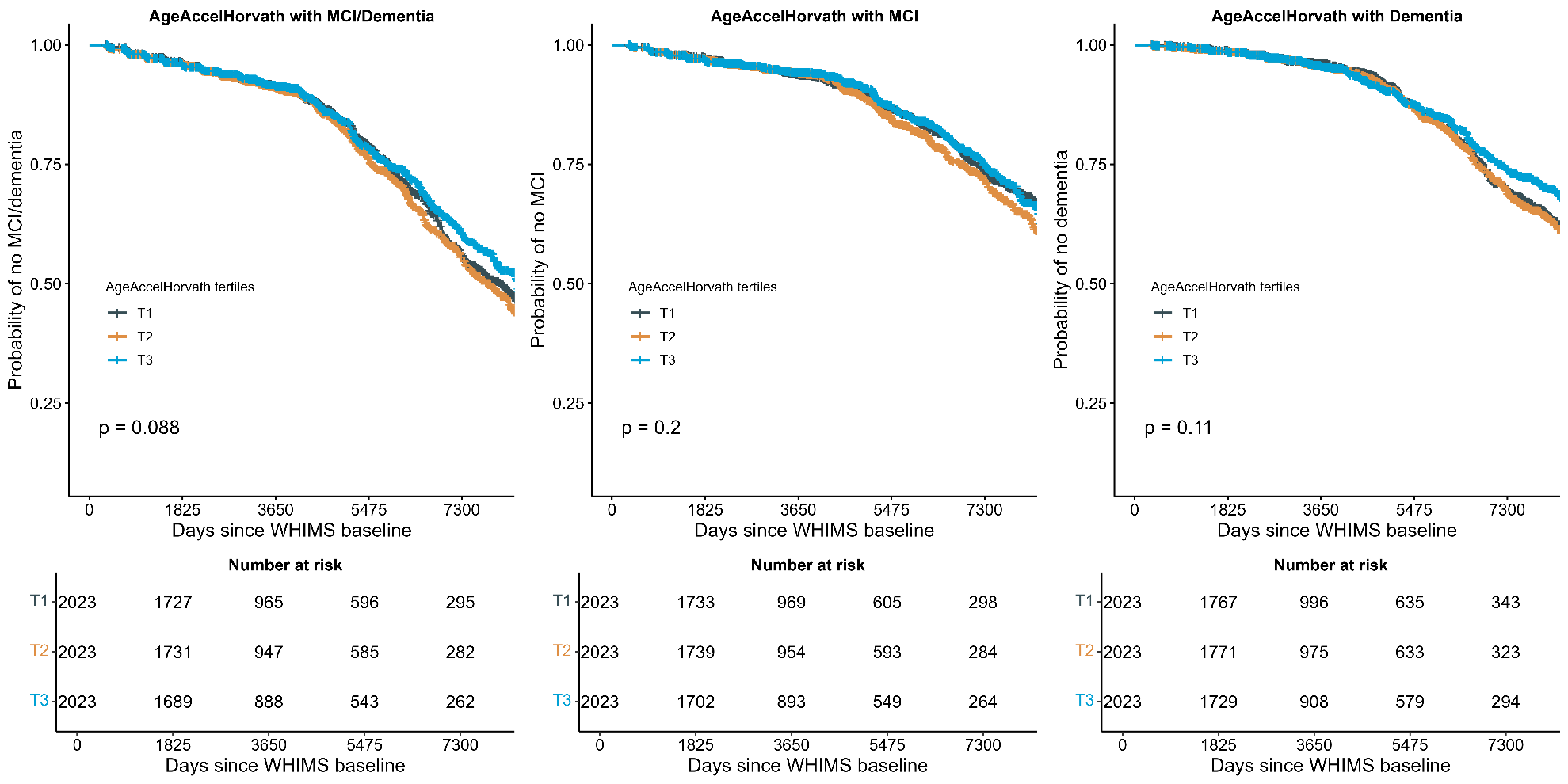
Abbreviations: MCI=mild cognitive impairment
p-values were calculated from a log rank test.

**eFigure 3. Kaplan Meier curves for mild cognitive impairment (MCI)/probable dementia, MCI, and probable dementia across tertiles of AgeAccelHannum in the Women’s Health Initiative Memory Study (WHIMS; n=6069) 1995-2021.**

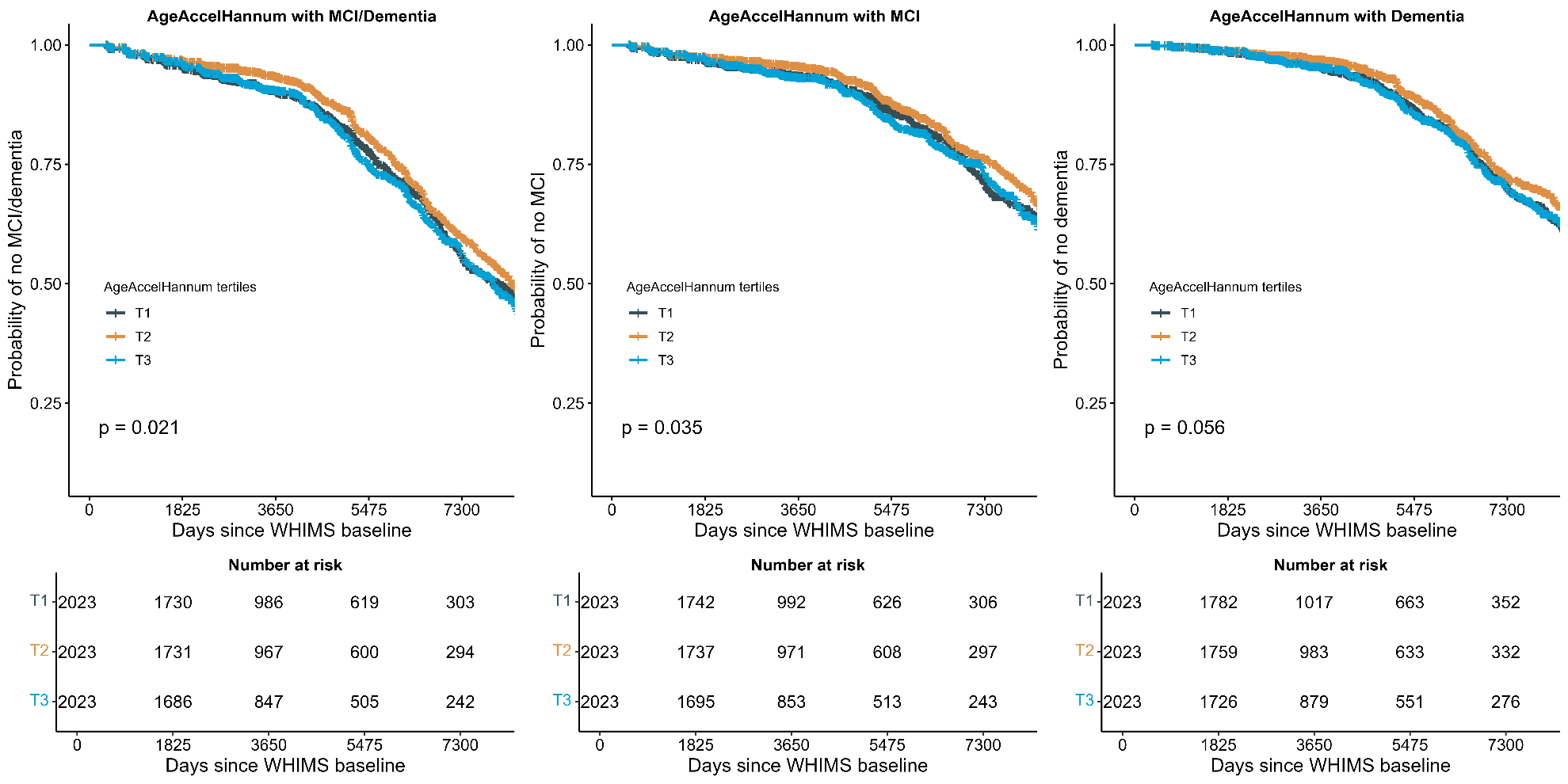
Abbreviations: MCI=mild cognitive impairment
p-values were calculated from a log rank test.

**eFigure 4. Kaplan Meier curves for mild cognitive impairment (MCI)/probable dementia, MCI, and probable dementia across tertiles of AgeAccelPheno in the Women’s Health Initiative Memory Study (WHIMS; n=6069) 1995-2021.**
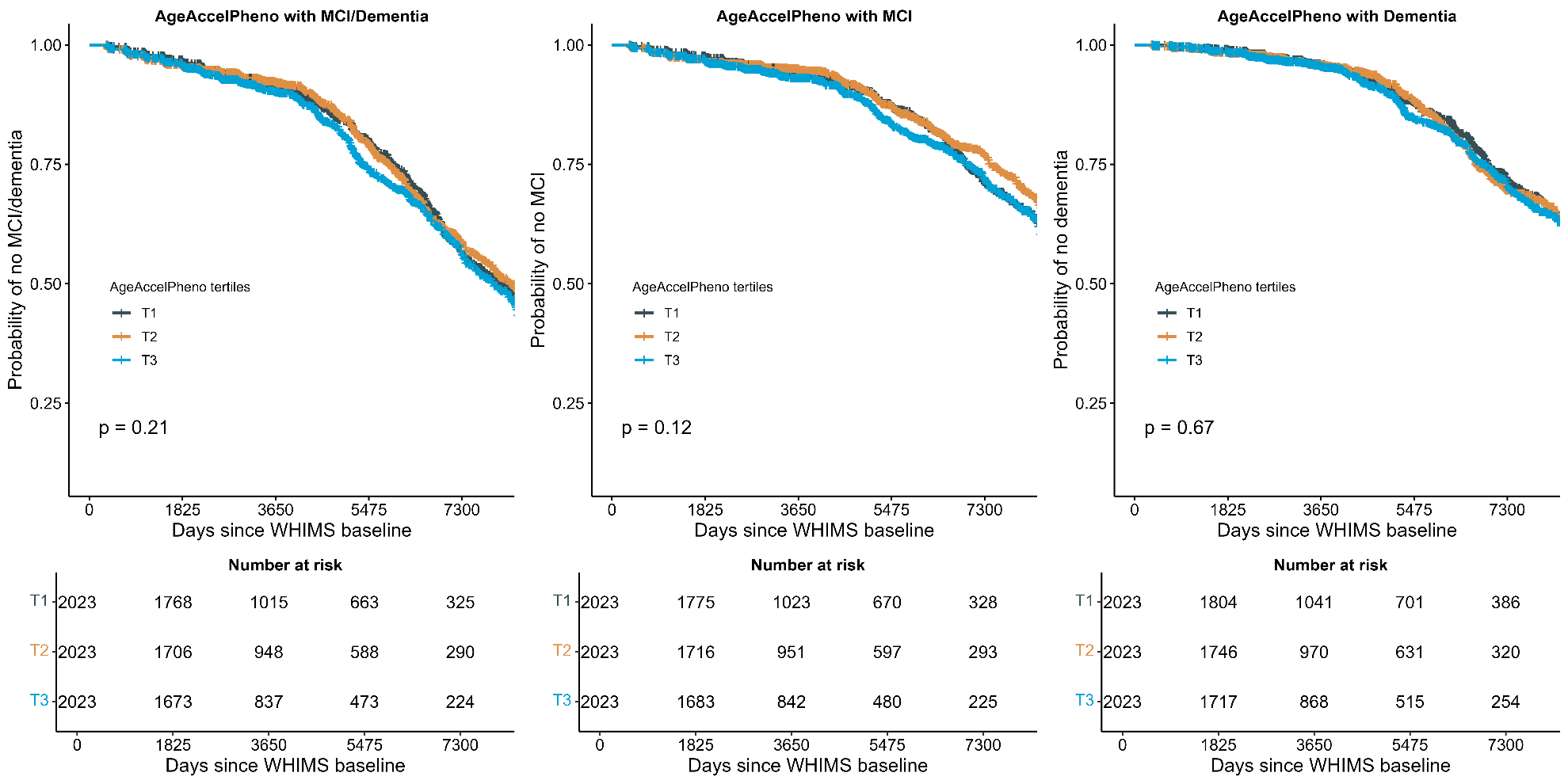
Abbreviations: MCI=mild cognitive impairment
p-values were calculated from a log rank test.

**eFigure 5. Kaplan Meier curves for mild cognitive impairment (MCI)/probable dementia, MCI, and probable dementia across tertiles of AgeAccelGrim2 in the Women’s Health Initiative Memory Study (WHIMS; n=6069) 1995-2021.**
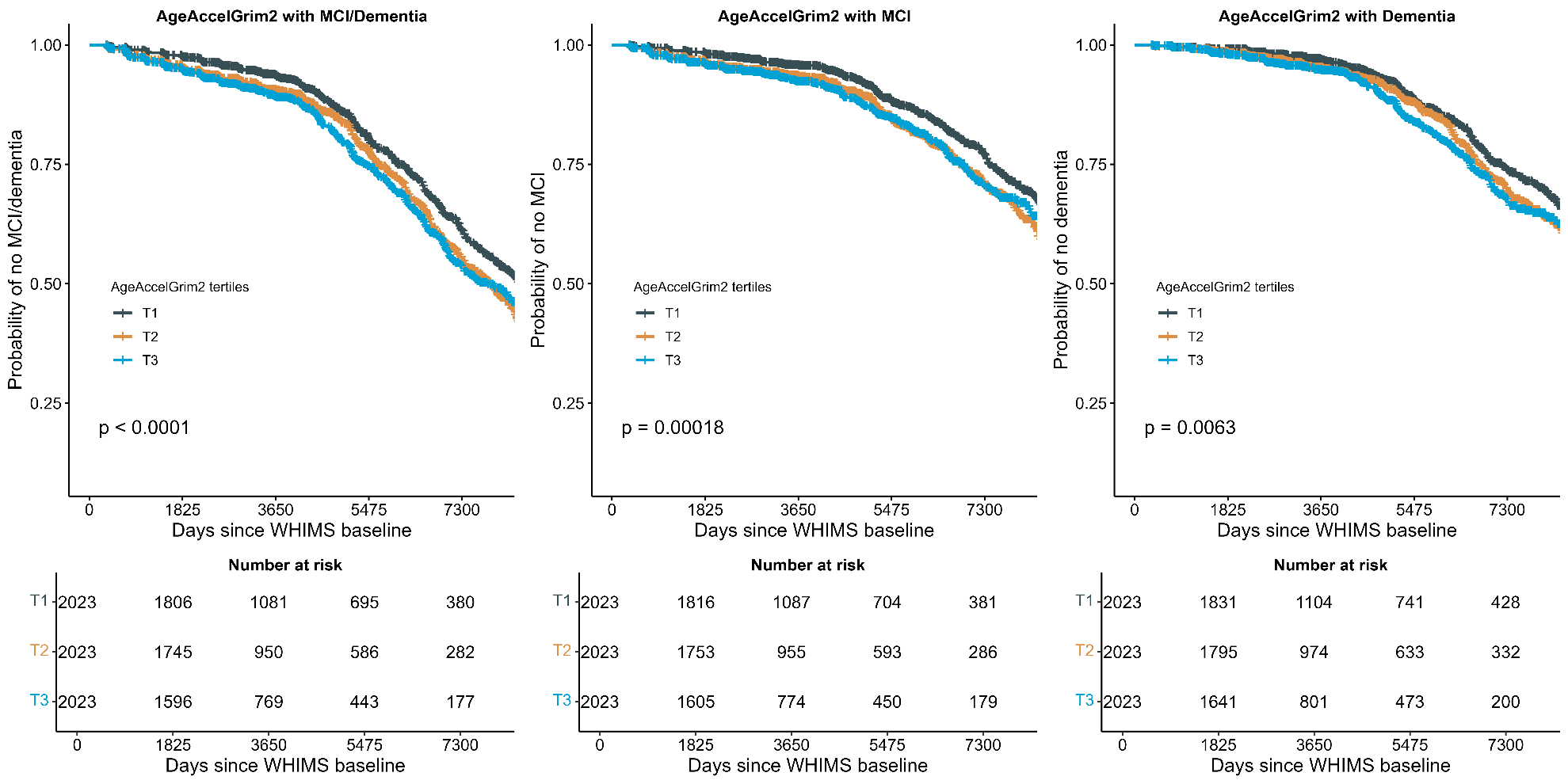
Abbreviations: MCI=mild cognitive impairment
p-values were calculated from a log rank test.

**eFigure 6. Kaplan Meier curves for mild cognitive impairment (MCI)/probable dementia, MCI, and probable dementia across tertiles of DunedinPACE in the Women’s Health Initiative Memory Study (WHIMS; n=6069) 1995-2021.**
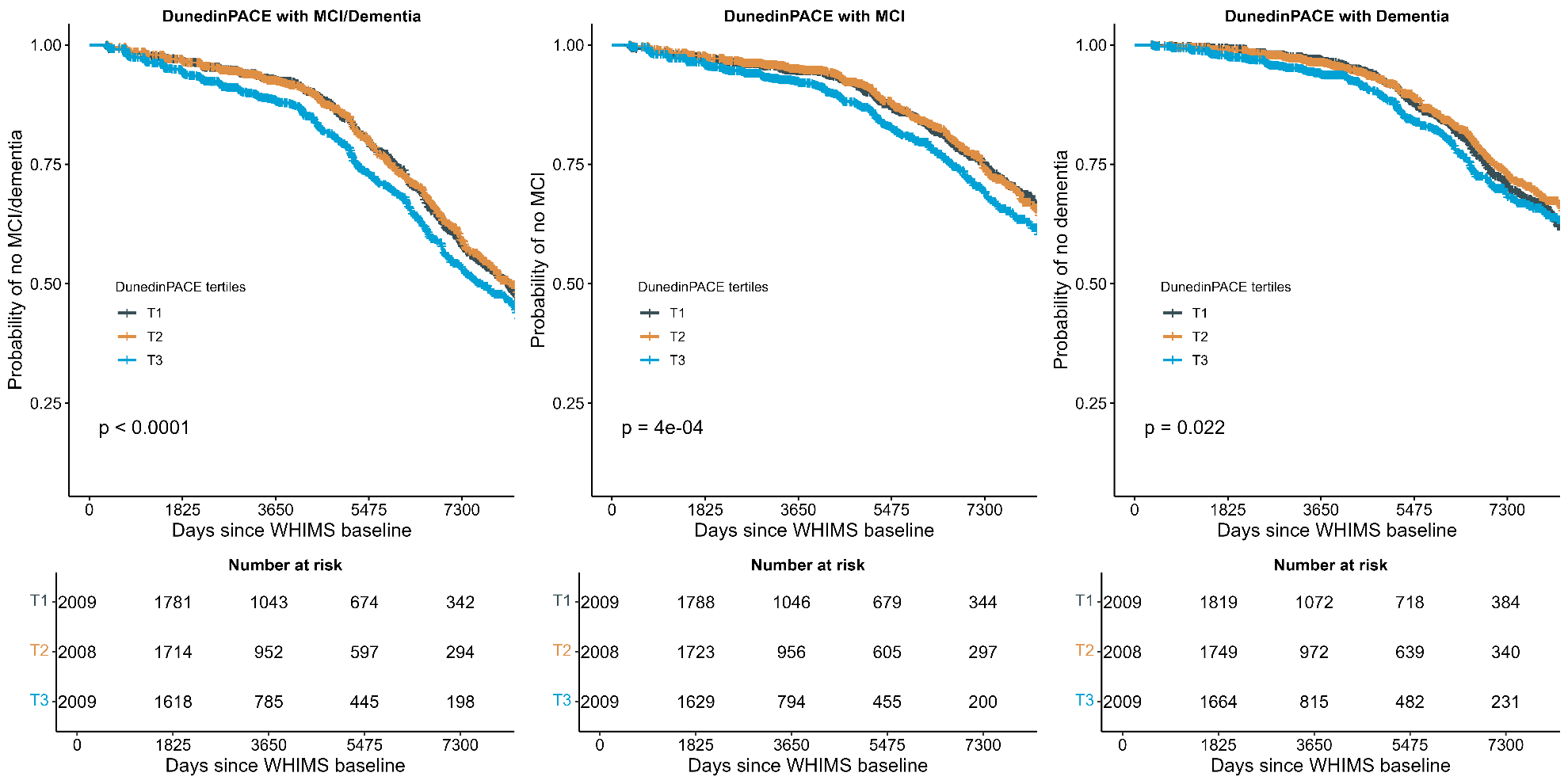
Abbreviations: MCI=mild cognitive impairment
p-values were calculated from a log rank test.

**eFigure 7. Associations of epigenetic clocks and AgeAccelGrim2 components with incident mild cognitive impairment (MCI)/probable dementia, MCI, and probable dementia in the Women’s Health Initiative Memory Study (WHIMS; n=6069) 1995-2021.**
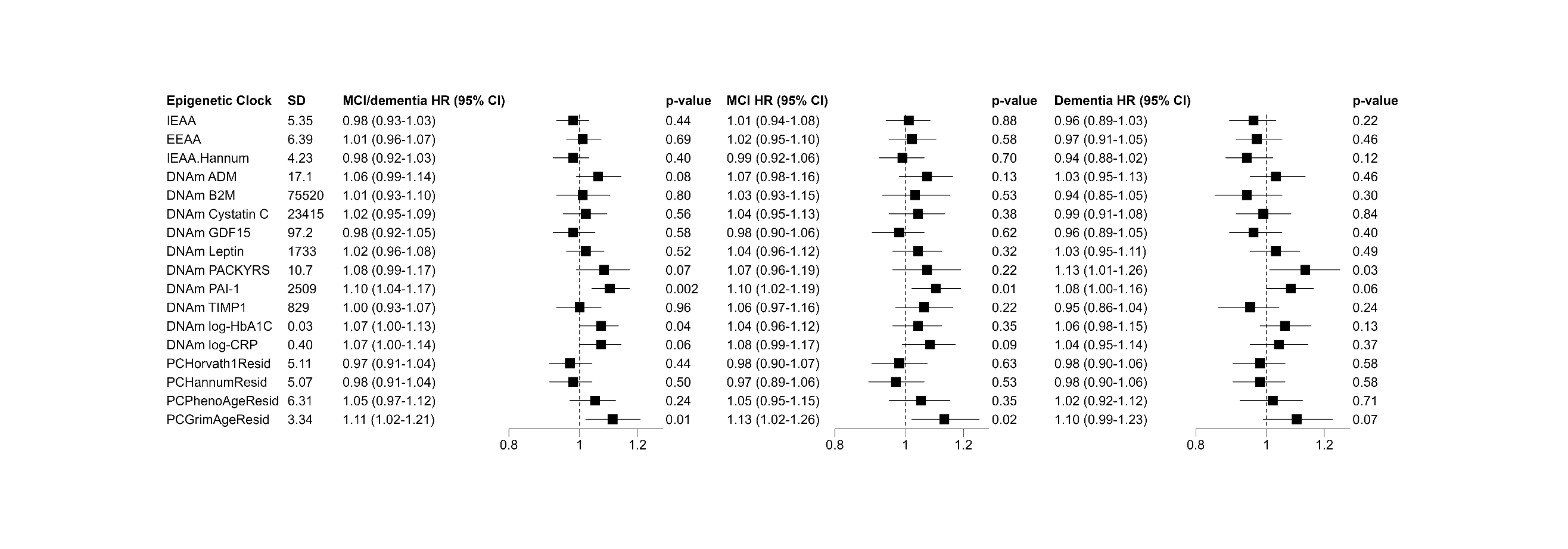

Abbreviations: SD=standard deviation; MCI=mild cognitive impairment; HR=hazard ratio; CI=confidence interval
There were 1307 incident MCI/dementia events, 799 incident MCI events, and 786 incident probable dementia events in an analytic sample of 6069 for analyses with AgeAccelHorvath, AgeAccelHannum, IEAA, EEAA, IEAA.Hannum, AgeAccelPheno, and AgeAccelGrim2.

There were 1301 incident MCI/dementia events, 797 incident MCI events, and 782 incident probable dementia events in an analytic sample of 6026 for analyses with PCHorvath1Resid, PCHannumResid, PCPhenoAgeResid, and PCGrimAgeResid.

The sum of separate incident MCI and probable dementia events is higher than the 1307 incident MCI/dementia events because participants with incident MCI later had incident probable dementia.

Models adjusted for age, education, smoking, race, ethnicity, hormone therapy trial arm, total energy expenditure from recreational physical activity, body mass index, and white blood cell counts (CD8T, CD4T, natural killer cells, B cells, monocytes, and neutrophils).

Models with IEAA, EEAA, and IEAA.Hannum did not include white blood cell counts.

Missing covariate data were imputed using multiple imputation by chained equations with the R *mice* package.

**eFigure 8. Associations of epigenetic clocks and AgeAccelGrim2 components with incident mild cognitive impairment (MCI)/probable dementia, MCI, and probable dementia in the Women’s Health Initiative Memory Study after excluding data from 79 women who had incident MCI/probable dementia during the first 2 years of follow-up (WHIMS; n=5990) 1995-2021.**

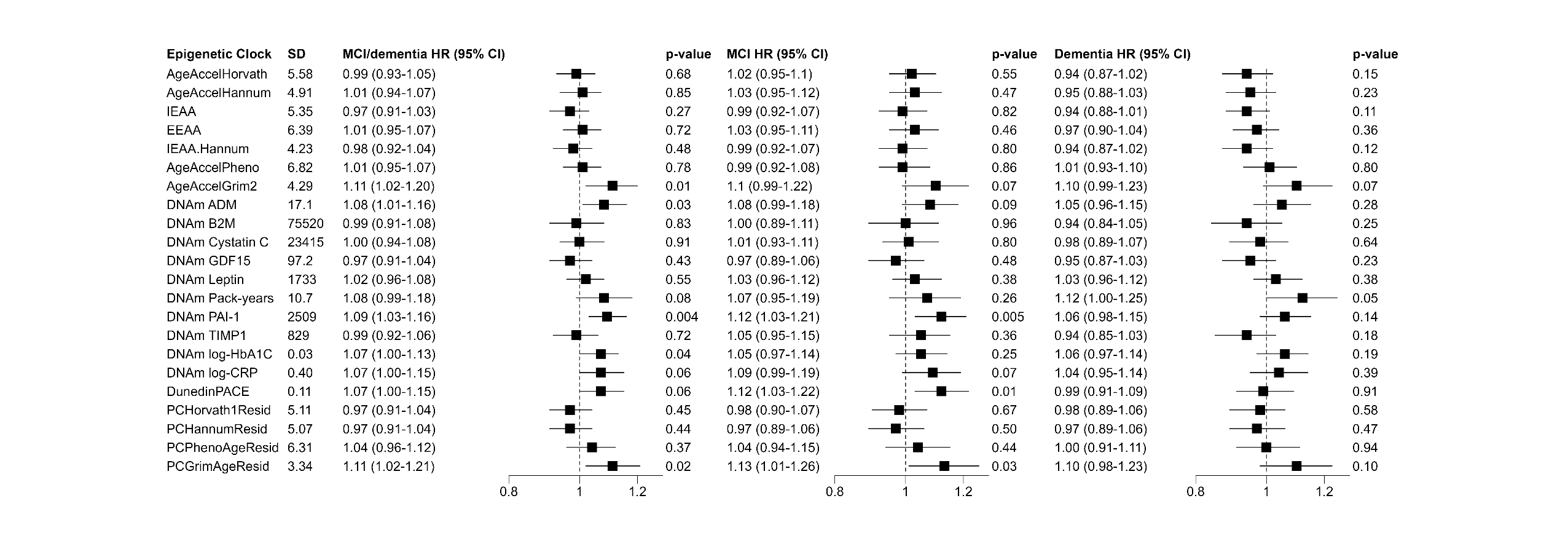

Abbreviations: SD=standard deviation; MCI=mild cognitive impairment; HR=hazard ratio; CI=confidence interval
There were 1228 incident MCI/probable dementia events, 740 incident MCI events, and 746 incident probable dementia events in an analytic sample of 5990 for analyses with AgeAccelHorvath, AgeAccelHannum, IEAA, EEAA, IEAA.Hannum, AgeAccelPheno, and AgeAccelGrim2.
There were 1222 incident MCI/probable dementia events, 738 incident MCI events, and 742 incident probable dementia events in an analytic sample of 5947 for analyses with DunedinPACE, PCHorvath1Resid, PCHannumResid, PCPhenoAgeResid, and PCGrimAgeResid.

The sum of separate incident MCI and probable dementia events is higher than the 1307 incident MCI/dementia events because participants with incident MCI later had incident probable dementia.

Models adjusted for age, education, smoking, race/ethnicity, hormone therapy trial arm, total energy expenditure from recreational physical activity, body mass index, and white blood cell counts (CD8T, CD4T, natural killer cells, B cells, monocytes, and neutrophils).
Models with IEAA, EEAA, and IEAA.Hannum did not include white blood cell counts.

Missing covariate data were imputed using multiple imputation by chained equations with the R *mice* package.

### **eTable 1. Baseline sociodemographic, behavior, and health characteristics by quartiles of DunedinPACE in the Women’s Health Initiative Memory Study (n=6026)**

|  |  | DunedinPACE quartiles | | | |  |
| --- | --- | --- | --- | --- | --- | --- |
| Characteristics | Total | Q1 (n=1507): [0.51-, 0.98] | Q2 (n=1506): (0.98, 1.05] | Q3 (n=1506): (1.05, 1.12] | Q4 (n=1507): (1.12, 1.64) | p-value |
|  | No. (%) | No. (%) | No. (%) | No. (%) | No. (%) |  |
| Age, *mean (sd)* | 70.0 (3.8) | 69.8 (3.7) | 70.1 (3.8) | 70.3 (3.9) | 69.9 (3.8) | 0.009 |
| Highest education level, *n (%)* |  |  |  |  |  | <0.001 |
| Less than high school/GED | 421 (7) | 72 (4.8) | 94 (6.3) | 113 (7.5) | 141 (9.4) |  |
| High school/GED | 1306 (21.6) | 328 (21.8) | 331 (22.1) | 338 (22.5) | 304 (20.3) |  |
| Some college | 2414 (39.9) | 544 (36.2) | 590 (39.3) | 621 (41.4) | 637 (42.5) |  |
| College graduate or higher | 1906 (31.5) | 559 (37.2) | 486 (32.4) | 428 (28.5) | 418 (27.9) |  |
| Hormone trial arm |  |  |  |  |  | <0.001 |
| Estrogen alone intervention | 1152 (19) | 249 (16.5) | 291 (19.3) | 293 (19.5) | 308 (20.4) |  |
| Estrogen placebo | 1163 (19.2) | 238 (15.8) | 262 (17.4) | 311 (20.7) | 348 (23.1) |  |
| Estrogen plus progestin intervention | 1854 (30.5) | 490 (32.5) | 482 (32) | 457 (30.3) | 406 (26.9) |  |
| Estrogen plus progestin placebo | 1900 (31.3) | 530 (35.2) | 471 (31.3) | 445 (29.5) | 445 (29.5) |  |
| Race, *n (%)* |  |  |  |  |  | <0.001 |
| American Indian or Alaskan Native | 13 (0.2) | 2 (0.1) | 4 (0.3) | 2 (0.1) | 5 (0.3) |  |
| Asian | 106 (1.7) | 16 (1.1) | 20 (1.3) | 42 (2.8) | 28 (1.9) |  |
| Native Hawaiian or other Pacific Islander | 8 (0.1) | 0 (0) | 3 (0.2) | 2 (0.1) | 3 (0.2) |  |
| Black | 417 (6.9) | 56 (3.7) | 79 (5.2) | 97 (6.4) | 181 (12) |  |
| White | 5403 (89) | 1417 (94) | 1377 (91.4) | 1324 (87.9) | 1247 (82.7) |  |
| More than 1 race | 68 (1.1) | 11 (0.7) | 14 (0.9) | 25 (1.7) | 18 (1.2) |  |
| Uknown or not reported | 54 (0.9) | 5 (0.3) | 9 (0.6) | 14 (0.9) | 25 (1.7) |  |
| Ethnicity, *n (%)* |  |  |  |  |  | <0.001 |
| Not Hispanic or Latina | 5890 (97.4) | 1484 (98.8) | 1470 (97.7) | 1457 (97.1) | 1437 (95.7) |  |
| Hispanic or Latina | 160 (2.6) | 18 (1.2) | 34 (2.3) | 43 (2.9) | 64 (4.3) |  |
| Unknown or not reported | 19 (0.3) | 5 (0.3) | 2 (0.1) | 6 (0.4) | 6 (0.4) |  |
| **Health behaviors/status** |  |  |  |  |  |  |
| Smoking status, *n (%)* |  |  |  |  |  | <0.001 |
| Never smoked | 3187 (53.3) | 927 (62.4) | 867 (58) | 774 (52.3) | 594 (40.2) |  |
| Past smoker | 2379 (39.8) | 528 (35.6) | 569 (38.1) | 612 (41.4) | 654 (44.3) |  |
| Current smoker | 412 (6.9) | 30 (2) | 59 (3.9) | 93 (6.3) | 229 (15.5) |  |
| Body mass index; kg/m^2^, *mean (SD)* | 28.6 (5.7) | 26.8 (5.1) | 28.0 (5.5) | 28.9 (5.5) | 30.6 (6.0) | <0.001 |
| Total energy expenditure from recreational physical activity; MET-hours/week, *mean (SD)* | 11.4 (13.2) | 13.8 (14.5) | 11.7 (13.1) | 10.8 (12.6) | 9.1 (12.2) | <0.001 |
| *APOE ε4* carriage | 1306 (25.3) | 361 (26.4) | 317 (23.9) | 311 (24.7) | 303 (25.8) | 0.46 |

Intervals are expressed as (a, b], where a < x ≤ b; the Q1 interval includes the minimum value

### **eTable 2.** **Baseline sociodemographic, behavior, and health characteristics by quartiles of AgeAccelPheno in the Women’s Health Initiative Memory Study (n=6069).**

|  |  | AgeAccelPheno quartiles | | | |  |
| --- | --- | --- | --- | --- | --- | --- |
| Characteristics | Total | Q1 (n=1507):  [-25.6, -5.25] | Q2 (n=1506): (-5.25, -1.03] | Q3 (n=1506): (-1.03, 3.28] | Q4 (n=1507): (3.28, 34.2] | p-value |
|  | No. (%) | No. (%) | No. (%) | No. (%) | No. (%) |  |
| Age, *mean (sd)* | 70.0 (3.8) | 70.0 (3.8) | 69.8 (3.7) | 70.2 (3.8) | 70.2 (3.9) | 0.003 |
| Highest education level, *n (%)* |  |  |  |  |  | 0.007 |
| Less than high school/GED | 421 (7) | 96 (6.4) | 91 (6) | 102 (6.7) | 132 (8.7) |  |
| High school/GED | 1306 (21.6) | 306 (20.3) | 337 (22.3) | 339 (22.4) | 324 (21.4) |  |
| Some college | 2414 (39.9) | 581 (38.5) | 596 (39.4) | 622 (41.1) | 615 (40.6) |  |
| College graduate or higher | 1906 (31.5) | 526 (34.9) | 488 (32.3) | 449 (29.7) | 443 (29.3) |  |
| Hormone trial arm |  |  |  |  |  | 0.40 |
| Estrogen alone intervention | 1152 (19) | 273 (18) | 296 (19.5) | 275 (18.1) | 308 (20.3) |  |
| Estrogen alone placebo | 1163 (19.2) | 280 (18.4) | 278 (18.3) | 307 (20.2) | 298 (19.6) |  |
| Estrogen plus progestin intervention | 1854 (30.5) | 470 (31) | 467 (30.8) | 447 (29.5) | 470 (31) |  |
| Estrogen plus progestin placebo | 1900 (31.3) | 495 (32.6) | 476 (31.4) | 488 (32.2) | 441 (29.1) |  |
| Race, *n (%)* |  |  |  |  |  | <0.001 |
| American Indian or Alaskan Native | 13 (0.2) | 1 (0.1) | 5 (0.3) | 4 (0.3) | 3 (0.2) |  |
| Asian | 106 (1.7) | 23 (1.5) | 24 (1.6) | 27 (1.8) | 32 (2.1) |  |
| Native Hawaiian or other Pacific Islander | 8 (0.1) | 2 (0.1) | 2 (0.1) | 2 (0.1) | 2 (0.1) |  |
| Black | 417 (6.9) | 162 (10.7) | 93 (6.1) | 86 (5.7) | 76 (5) |  |
| White | 5403 (89) | 1310 (86.3) | 1364 (89.9) | 1360 (89.7) | 1369 (90.2) |  |
| More than 1 race | 68 (1.1) | 12 (0.8) | 18 (1.2) | 23 (1.5) | 15 (1) |  |
| Uknown or not reported | 54 (0.9) | 8 (0.5) | 11 (0.7) | 15 (1) | 20 (1.3) |  |
| Ethnicity, *n (%)* |  |  |  |  |  | 0.45 |
| Not Hispanic or Latina | 5890 (97.1) | 1485 (97.8) | 1466 (96.6) | 1473 (97.1) | 1466 (96.6) |  |
| Hispanic or Latina | 160 (2.6) | 30 (2) | 46 (3) | 40 (2.6) | 44 (2.9) |  |
| Unknown or not reported | 19 (0.3) | 3 (0.2) | 5 (0.3) | 4 (0.3) | 7 (0.5) |  |
| **Health behaviors/status** |  |  |  |  |  |  |
| Smoking status, *n (%)* |  |  |  |  |  | 0.001 |
| Never smoked | 3187 (53.3) | 836 (55.8) | 831 (55.8) | 771 (51.6) | 749 (50.1) |  |
| Past smoker | 2379 (39.8) | 582 (38.8) | 567 (38.1) | 608 (40.7) | 622 (41.6) |  |
| Current smoker | 412 (6.9) | 81 (5.4) | 92 (6.2) | 114 (7.6) | 125 (8.4) |  |
| Body mass index; kg/m^2^, *mean (SD)* | 28.6 (5.7) | 27.9 (5.4) | 28.5 (5.7) | 28.7 (5.6) | 29.1 (6.0) | <0.001 |
| Total energy expenditure from recreational physical activity; MET-hours/week, *mean (SD)* | 11.4 (13.2) | 12.5 (13.6) | 11.4 (13.0) | 10.8 (13.1) | 10.7 (13.1) | <0.001 |
| *APOE ε4* carriage | 1306 (25.3) | 301 (23.8) | 343 (26.2) | 336 (26) | 326 (25.1) | 0.49 |

Intervals are expressed as (a, b], where a < x ≤ b; the Q1 interval includes the minimum value

### **eTable 3. Baseline sociodemographic, behavioral, and health characteristics of 6069 Women’s Health Initiative Memory Study women included in the analytic sample versus those excluded (n=1410)**

| Characteristics | Total | In analytic sample (n=6069) | Excluded (n=1410) | p-value |
| --- | --- | --- | --- | --- |
|  | No. (%) | No. (%) | No. (%) |  |
| Age, *mean (sd)* | 70.12 (3.84) | 70.0 (3.8) | 70.52, (3.95) | <0.001 |
| Highest education level, *n (%)* |  |  |  | <0.001 |
| Less than high school/GED | 576 (7.7) | 421 (7) | 155 (11) |  |
| High school/GED | 1647 (22.1) | 1306 (21.6) | 341 (24.2) |  |
| Some college | 3002 (40.3) | 2414 (39.9) | 588 (41.7) |  |
| College graduate or higher | 2232 (29.9) | 1906 (31.5) | 326 (23.1) |  |
| Hormone trial arm |  |  |  | <0.001 |
| Estrogen alone intervention | 1469 (19.6) | 1152 (19) | 317 (22.5) |  |
| Estrogen alone placebo | 1484 (19.8) | 1163 (19.2) | 321 (22.8) |  |
| Estrogen plus progestin intervention | 2224 (29.7) | 1854 (30.5) | 370 (26.2) |  |
| Estrogen plus progestin placebo | 2302 (30.8) | 1900 (31.3) | 402 (28.5) |  |
| Race, *n (%)* |  |  |  | 0.23 |
| American Indian or Alaskan Native | 17 (0.2) | 13 (0.2) | 4 (0.3) |  |
| Asian | 127 (1.7) | 106 (1.7) | 21 (1.5) |  |
| Native Hawaiian or other Pacific Islander | 8 (0.1) | 8 (0.1) | 0 (0) |  |
| Black | 523 (7) | 417 (6.9) | 106 (7.5) |  |
| White | 6649 (88.9) | 5403 (89) | 1246 (88.4) |  |
| More than 1 race | 80 (1.1) | 68 (1.1) | 12 (0.9) |  |
| Uknown or not reported | 75 (1) | 54 (0.9) | 21 (1.5) |  |
| Ethnicity, *n (%)* |  |  |  | <0.001 |
| Not Hispanic or Latina | 7233 (96.7) | 5890 (97.4) | 1343 (95.2) |  |
| Hispanic or Latina | 214 (2.9) | 160 (2.6) | 54 (3.8) |  |
| **Health behaviors/status** |  |  |  |  |
| Smoking status, *n (%)* |  |  |  | 0.13 |
| Never smoked | 3909 (53.1) | 3187 (53.3) | 722 (51.9) |  |
| Past smoker | 2930 (39.8) | 2379 (39.8) | 551 (39.6) |  |
| Current smoker | 529 (7.2) | 412 (6.9) | 117 (8.4) |  |
| Body mass index; kg/m^2^, *mean (SD)* | 28.52 (5.7) | 28.6 (5.7) | 28.41, (5.74) | 0.42 |
| Total energy expenditure from recreational physical activity; MET-hours/week, *mean (SD)* | 11.25 (13.29) | 11.4 (13.2) | 10.82, (13.6) | 0.18 |
| *APOE ε4* carriage | 1480 (25.2) | 1306 (25.3) | 174 (24.9) | 0.86 |

**eTable 4. Associations of epigenetic clocks with incident mild cognitive impairment (MCI)/probable dementia, MCI, and probable dementia in the Women’s Health Initiative Memory Study (WHIMS; n=6069) 1995-2021.**

|  | **AgeAccelHorvath (years)** |  |  |  |
| --- | --- | --- | --- | --- |
|  | HR (95% CI) for 1-SD increment (5.58) | P-value^a^ | n | n events |
| **MCI/Dementia** |  |  |  |  |
| Model 1 | 0.98 (0.93-1.03) | 0.47 | 6069 | 1307 |
| Model 2 | 1.01 (0.95-1.06) | 0.82 | 6069 | 1307 |
| Model 3 | 1.00 (0.95-1.06) | 0.98 | 6069 | 1307 |
| Model 4 | 1.00 (0.94-1.06) | 0.98 | 6069 | 1307 |
| **MCI** |  |  |  |  |
| Model 1 | 1.00 (0.93-1.07) | 0.98 | 6069 | 799 |
| Model 2 | 1.04 (0.97-1.12) | 0.22 | 6069 | 799 |
| Model 3 | 1.03 (0.96-1.10) | 0.42 | 6069 | 799 |
| Model 4 | 1.04 (0.96-1.11) | 0.35 | 6069 | 799 |
| **Dementia** |  |  |  |  |
| Model 1 | 0.96 (0.89-1.03) | 0.23 | 6069 | 786 |
| Model 2 | 0.97 (0.90-1.04) | 0.35 | 6069 | 786 |
| Model 3 | 0.96 (0.90-1.04) | 0.32 | 6069 | 786 |
| Model 4 | 0.96 (0.89-1.04) | 0.29 | 6069 | 786 |
|  | **AgeAccelHannum (years)** |  |  |  |
|  | HR (95% CI) for 1-SD increment (4.91) | P-value^a^ | n | n events |
| **MCI/Dementia** |  |  |  |  |
| Model 1 | 0.98 (0.93-1.04) | 0.59 | 6069 | 1307 |
| Model 2 | 1.02 (0.96-1.07) | 0.60 | 6069 | 1307 |
| Model 3 | 1.01 (0.96-1.07) | 0.69 | 6069 | 1307 |
| Model 4 | 1.01 (0.95-1.07) | 0.82 | 6069 | 1307 |
| **MCI** |  |  |  |  |
| Model 1 | 0.98 (0.92-1.05) | 0.63 | 6069 | 799 |
| Model 2 | 1.03 (0.96-1.11) | 0.37 | 6069 | 799 |
| Model 3 | 1.03 (0.96-1.10) | 0.48 | 6069 | 799 |
| Model 4 | 1.03 (0.95-1.11) | 0.51 | 6069 | 799 |
| **Dementia** |  |  |  |  |
| Model 1 | 0.96 (0.90-1.03) | 0.27 | 6069 | 786 |
| Model 2 | 0.97 (0.90-1.04) | 0.40 | 6069 | 786 |
| Model 3 | 0.97 (0.90-1.04) | 0.39 | 6069 | 786 |
| Model 4 | 0.96 (0.88-1.03) | 0.27 | 6069 | 786 |
|  | **IEAA (years)** |  |  |  |
|  | HR (95% CI) for 1-SD increment (5.35) | P-value^a^ | n | n events |
| **MCI/Dementia** |  |  |  |  |
| Model 1 | 0.96 (0.91-1.02) | 0.20 | 6069 | 1307 |
| Model 2 | 0.98 (0.93-1.04) | 0.53 | 6069 | 1307 |
| Model 3 | 0.98 (0.93-1.03) | 0.44 | 6069 | 1307 |
| Model 4 | 0.98 (0.93-1.03) | 0.44 | 6069 | 1307 |
| **MCI** |  |  |  |  |
| Model 1 | 0.99 (0.92-1.06) | 0.70 | 6069 | 799 |
| Model 2 | 1.01 (0.95-1.09) | 0.70 | 6069 | 799 |
| Model 3 | 1.01 (0.94-1.08) | 0.88 | 6069 | 799 |
| Model 4 | 1.01 (0.94-1.08) | 0.88 | 6069 | 799 |
| **Dementia** |  |  |  |  |
| Model 1 | 0.95 (0.88-1.02) | 0.15 | 6069 | 786 |
| Model 2 | 0.96 (0.89-1.03) | 0.23 | 6069 | 786 |
| Model 3 | 0.96 (0.89-1.03) | 0.22 | 6069 | 786 |
| Model 4 | 0.96 (0.89-1.03) | 0.22 | 6069 | 786 |
|  | **EEAA (years)** |  |  |  |
|  | HR (95% CI) for 1-SD increment (6.39) | P-value^a^ | n | n events |
| **MCI/Dementia** |  |  |  |  |
| Model 1 | 0.99 (0.93-1.04) | 0.63 | 6069 | 1307 |
| Model 2 | 1.02 (0.96-1.07) | 0.58 | 6069 | 1307 |
| Model 3 | 1.01 (0.96-1.07) | 0.69 | 6069 | 1307 |
| Model 4 | 1.01 (0.96-1.07) | 0.69 | 6069 | 1307 |
| **MCI** |  |  |  |  |
| Model 1 | 0.98 (0.91-1.05) | 0.54 | 6069 | 799 |
| Model 2 | 1.03 (0.96-1.10) | 0.44 | 6069 | 799 |
| Model 3 | 1.02 (0.95-1.10) | 0.58 | 6069 | 799 |
| Model 4 | 1.02 (0.95-1.10) | 0.58 | 6069 | 799 |
| **Dementia** |  |  |  |  |
| Model 1 | 0.97 (0.90-1.04) | 0.36 | 6069 | 786 |
| Model 2 | 0.97 (0.91-1.05) | 0.48 | 6069 | 786 |
| Model 3 | 0.97 (0.91-1.05) | 0.46 | 6069 | 786 |
| Model 4 | 0.97 (0.91-1.05) | 0.46 | 6069 | 786 |
|  | **IEAA.Hannum (years)** |  |  |  |
|  | HR (95% CI) for 1-SD increment (4.23) | P-value^a^ | n | n events |
| **MCI/Dementia** |  |  |  |  |
| Model 1 | 0.96 (0.91-1.02) | 0.15 | 6069 | 1307 |
| Model 2 | 0.98 (0.92-1.03) | 0.43 | 6069 | 1307 |
| Model 3 | 0.98 (0.92-1.03) | 0.40 | 6069 | 1307 |
| Model 4 | 0.98 (0.92-1.03) | 0.40 | 6069 | 1307 |
| **MCI** |  |  |  |  |
| Model 1 | 0.96 (0.89-1.03) | 0.25 | 6069 | 799 |
| Model 2 | 0.99 (0.92-1.06) | 0.75 | 6069 | 799 |
| Model 3 | 0.99 (0.92-1.06) | 0.70 | 6069 | 799 |
| Model 4 | 0.99 (0.92-1.06) | 0.70 | 6069 | 799 |
| **Dementia** |  |  |  |  |
| Model 1 | 0.94 (0.87-1.01) | 0.08 | 6069 | 786 |
| Model 2 | 0.94 (0.88-1.01) | 0.11 | 6069 | 786 |
| Model 3 | 0.94 (0.88-1.02) | 0.12 | 6069 | 786 |
| Model 4 | 0.94 (0.88-1.02) | 0.12 | 6069 | 786 |
|  | **AgeAccelPheno (years)** |  |  |  |
|  | HR (95% CI) for 1-SD increment (6.82) | P-value^a^ | n | n events |
| **MCI/ementia** |  |  |  |  |
| Model 1 | 1.01 (0.96-1.07) | 0.64 | 6069 | 1307 |
| Model 2 | 1.02 (0.97-1.08) | 0.42 | 6069 | 1307 |
| Model 3 | 1.02 (0.96-1.08) | 0.51 | 6069 | 1307 |
| Model 4 | 1.01 (0.95-1.07) | 0.74 | 6069 | 1307 |
| **MCI** |  |  |  |  |
| Model 1 | 0.99 (0.93-1.07) | 0.89 | 6069 | 799 |
| Model 2 | 1.01 (0.94-1.08) | 0.78 | 6069 | 799 |
| Model 3 | 1.00 (0.93-1.07) | 0.98 | 6069 | 799 |
| Model 4 | 0.99 (0.92-1.07) | 0.89 | 6069 | 799 |
| **Dementia** |  |  |  |  |
| Model 1 | 1.03 (0.96-1.10) | 0.44 | 6069 | 786 |
| Model 2 | 1.03 (0.96-1.11) | 0.37 | 6069 | 786 |
| Model 3 | 1.03 (0.96-1.11) | 0.41 | 6069 | 786 |
| Model 4 | 1.02 (0.94-1.10) | 0.62 | 6069 | 786 |
|  | **AgeAccelGrim2 (years)** |  |  |  |
|  | HR (95% CI) for 1-SD increment (4.29) | P-value^a^ | n | n events |
| **MCI/Dementia** |  |  |  |  |
| Model 1 | 1.14 (1.07-1.20) | <0.001 | 6069 | 1307 |
| Model 2 | 1.10 (1.04-1.17) | 0.001 | 6069 | 1307 |
| Model 3 | 1.11 (1.04-1.19) | 0.003 | 6069 | 1307 |
| Model 4 | 1.11 (1.02-1.20) | 0.01 | 6069 | 1307 |
| **MCI** |  |  |  |  |
| Model 1 | 1.13 (1.05-1.22) | 0.001 | 6069 | 799 |
| Model 2 | 1.07 (1.00-1.15) | 0.07 | 6069 | 799 |
| Model 3 | 1.08 (0.99-1.18) | 0.07 | 6069 | 799 |
| Model 4 | 1.10 (1.00-1.22) | 0.06 | 6069 | 799 |
| **Dementia** |  |  |  |  |
| Model 1 | 1.13 (1.05-1.22) | 0.001 | 6069 | 786 |
| Model 2 | 1.12 (1.04-1.21) | 0.004 | 6069 | 786 |
| Model 3 | 1.14 (1.04-1.25) | 0.005 | 6069 | 786 |
| Model 4 | 1.11 (1.00-1.23) | 0.05 | 6069 | 786 |
|  | **DunedinPACE (% faster pace of aging compared with the norm for midlife adults)** | | | |
|  | HR (95% CI) for 1-SD increment (0.11) | P-value^a^ | n | n events |
| **MCI/Dementia** |  |  |  |  |
| Model 1 | 1.11 (1.05-1.18) | <0.001 | 6026 | 1301 |
| Model 2 | 1.07 (1.01-1.14) | 0.02 | 6026 | 1301 |
| Model 3 | 1.08 (1.01-1.15) | 0.02 | 6026 | 1301 |
| Model 4 | 1.07 (1.01-1.15) | 0.03 | 6026 | 1301 |
| **MCI** |  |  |  |  |
| Model 1 | 1.16 (1.08-1.25) | <0.001 | 6026 | 797 |
| Model 2 | 1.09 (1.01-1.17) | 0.02 | 6026 | 797 |
| Model 3 | 1.10 (1.02-1.19) | 0.02 | 6026 | 797 |
| Model 4 | 1.11 (1.02-1.21) | 0.01 | 6026 | 797 |
| **Dementia** |  |  |  |  |
| Model 1 | 1.05 (0.97-1.13) | 0.22 | 6026 | 782 |
| Model 2 | 1.03 (0.96-1.11) | 0.42 | 6026 | 782 |
| Model 3 | 1.03 (0.95-1.12) | 0.44 | 6026 | 782 |
| Model 4 | 1.01 (0.93-1.10) | 0.82 | 6026 | 782 |
|  | **PCHorvath1Resid (years)** | | | |
|  | HR (95% CI) for 1-SD increment (5.11) | P-value^a^ | n | n events |
| **MCI/Dementia** |  |  |  |  |
| Model 1 | 0.96 (0.91-1.02) | 0.18 | 6026 | 1301 |
| Model 2 | 0.99 (0.94-1.05) | 0.77 | 6026 | 1301 |
| Model 3 | 0.99 (0.93-1.04) | 0.63 | 6026 | 1301 |
| Model 4 | 0.97 (0.91-1.04) | 0.44 | 6026 | 1301 |
| **MCI** |  |  |  |  |
| Model 1 | 0.93 (0.87-1.00) | 0.05 | 6026 | 797 |
| Model 2 | 0.98 (0.91-1.05) | 0.59 | 6026 | 797 |
| Model 3 | 0.97 (0.90-1.05) | 0.45 | 6026 | 797 |
| Model 4 | 0.98 (0.90-1.07) | 0.63 | 6026 | 797 |
| **Dementia** |  |  |  |  |
| Model 1 | 0.99 (0.93-1.07) | 0.87 | 6026 | 782 |
| Model 2 | 1.00 (0.93-1.07) | 0.99 | 6026 | 782 |
| Model 3 | 1.00 (0.93-1.08) | 0.97 | 6026 | 782 |
| Model 4 | 0.98 (0.9-1.06) | 0.58 | 6026 | 782 |
|  | **PCHannumResid (years)** |  |  |  |
|  | HR (95% CI) for 1-SD increment (5.07) | P-value^a^ | n | n events |
| **MCI/Dementia** |  |  |  |  |
| Model 1 | 0.98 (0.92-1.03) | 0.40 | 6026 | 1301 |
| Model 2 | 1.00 (0.94-1.05) | 0.90 | 6026 | 1301 |
| Model 3 | 0.99 (0.94-1.05) | 0.77 | 6026 | 1301 |
| Model 4 | 0.98 (0.91-1.04) | 0.50 | 6026 | 1301 |
| **MCI** |  |  |  |  |
| Model 1 | 0.95 (0.88-1.01) | 0.12 | 6026 | 797 |
| Model 2 | 0.98 (0.91-1.05) | 0.57 | 6026 | 797 |
| Model 3 | 0.97 (0.90-1.04) | 0.43 | 6026 | 797 |
| Model 4 | 0.97 (0.89-1.06) | 0.53 | 6026 | 797 |
| **Dementia** |  |  |  |  |
| Model 1 | 1.00 (0.93-1.07) | 0.99 | 6026 | 782 |
| Model 2 | 1.00 (0.94-1.08) | 0.91 | 6026 | 782 |
| Model 3 | 1.00 (0.93-1.08) | 0.92 | 6026 | 782 |
| Model 4 | 0.98 (0.90-1.06) | 0.58 | 6026 | 782 |
|  | **PCPhenoAgeResid (years)** |  |  |  |
|  | HR (95% CI) for 1-SD increment (6.31) | P-value^a^ | n | n events |
| **MCI/Dementia** |  |  |  |  |
| Model 1 | 1.07 (1.01-1.13) | 0.02 | 6026 | 1301 |
| Model 2 | 1.05 (0.99-1.11) | 0.09 | 6026 | 1301 |
| Model 3 | 1.04 (0.98-1.10) | 0.15 | 6026 | 1301 |
| Model 4 | 1.05 (0.97-1.12) | 0.24 | 6026 | 1301 |
| **MCI** |  |  |  |  |
| Model 1 | 1.07 (0.99-1.15) | 0.07 | 6026 | 797 |
| Model 2 | 1.04 (0.97-1.11) | 0.30 | 6026 | 797 |
| Model 3 | 1.03 (0.95-1.10) | 0.49 | 6026 | 797 |
| Model 4 | 1.05 (0.95-1.15) | 0.35 | 6026 | 797 |
| **Dementia** |  |  |  |  |
| Model 1 | 1.06 (0.99-1.14) | 0.12 | 6026 | 782 |
| Model 2 | 1.05 (0.98-1.13) | 0.17 | 6026 | 782 |
| Model 3 | 1.05 (0.97-1.13) | 0.21 | 6026 | 782 |
| Model 4 | 1.02 (0.92-1.12) | 0.71 | 6026 | 782 |
|  | **PCGrimAgeResid (years)** |  |  |  |
|  | HR (95% CI) for 1-SD increment (3.34) | P-value^a^ | n | n events |
| **MCI/Dementia** |  |  |  |  |
| Model 1 | 1.11 (1.04-1.17) | 0.001 | 6026 | 1301 |
| Model 2 | 1.09 (1.03-1.15) | 0.004 | 6026 | 1301 |
| Model 3 | 1.09 (1.02-1.17) | 0.008 | 6026 | 1301 |
| Model 4 | 1.11 (1.02-1.21) | 0.01 | 6026 | 1301 |
| **MCI** |  |  |  |  |
| Model 1 | 1.09 (1.02-1.18) | 0.02 | 6026 | 797 |
| Model 2 | 1.07 (0.99-1.15) | 0.09 | 6026 | 797 |
| Model 3 | 1.08 (0.99-1.17) | 0.08 | 6026 | 797 |
| Model 4 | 1.13 (1.02-1.26) | 0.02 | 6026 | 797 |
| **Dementia** |  |  |  |  |
| Model 1 | 1.12 (1.04-1.21) | 0.003 | 6026 | 782 |
| Model 2 | 1.11 (1.03-1.20) | 0.005 | 6026 | 782 |
| Model 3 | 1.13 (1.04-1.23) | 0.005 | 6026 | 782 |
| Model 4 | 1.10 (0.99-1.23) | 0.07 | 6026 | 782 |
|  | **DNAm ADM** |  |  |  |
|  | HR (95% CI) for 1-SD increment (17.1) | P-value^a^ | n | n events |
| **MCI/Dementia** |  |  |  |  |
| Model 1 | 1.06 (1.00-1.12) | 0.07 | 6069 | 1307 |
| Model 2 | 1.07 (1.01-1.14) | 0.02 | 6069 | 1307 |
| Model 3 | 1.07 (1.01-1.14) | 0.03 | 6069 | 1307 |
| Model 4 | 1.06 (0.99-1.14) | 0.08 | 6069 | 1307 |
| **MCI** |  |  |  |  |
| Model 1 | 1.05 (0.97-1.13) | 0.24 | 6069 | 799 |
| Model 2 | 1.08 (1.00-1.16) | 0.05 | 6069 | 799 |
| Model 3 | 1.07 (0.99-1.16) | 0.09 | 6069 | 799 |
| Model 4 | 1.07 (0.98-1.16) | 0.13 | 6069 | 799 |
| **Dementia** |  |  |  |  |
| Model 1 | 1.06 (0.98-1.14) | 0.16 | 6069 | 786 |
| Model 2 | 1.06 (0.98-1.15) | 0.13 | 6069 | 786 |
| Model 3 | 1.05 (0.97-1.14) | 0.19 | 6069 | 786 |
| Model 4^c^ | 1.03 (0.95-1.13) | 0.46 | 6069 | 786 |
|  | **DNAm B2M** |  |  |  |
|  | HR (95% CI) for 1-SD increment (75,520) | P-value^a^ | n | n events |
| **MCI/Dementia** |  |  |  |  |
| Model 1 | 1.06 (0.98-1.14) | 0.14 | 6069 | 1307 |
| Model 2 | 1.04 (0.96-1.12) | 0.32 | 6069 | 1307 |
| Model 3 | 1.02 (0.95-1.10) | 0.59 | 6069 | 1307 |
| Model 4 | 1.01 (0.93-1.10) | 0.80 | 6069 | 1307 |
| **MCI** |  |  |  |  |
| Model 1 | 1.08 (0.98-1.18) | 0.12 | 6069 | 799 |
| Model 2 | 1.05 (0.95-1.15) | 0.32 | 6069 | 799 |
| Model 3 | 1.02 (0.93-1.12) | 0.67 | 6069 | 799 |
| Model 4 | 1.03 (0.93-1.15) | 0.53 | 6069 | 799 |
| **Dementia** |  |  |  |  |
| Model 1 | 1.01 (0.92-1.11) | 0.85 | 6069 | 786 |
| Model 2 | 1.00 (0.91-1.10) | 0.97 | 6069 | 786 |
| Model 3 | 0.98 (0.89-1.09) | 0.76 | 6069 | 786 |
| Model 4 | 0.94 (0.85-1.05) | 0.30 | 6069 | 786 |
|  | **DNAm Cystatin C** |  |  |  |
|  | HR (95% CI) for 1-SD increment (23,415) | P-value^a^ | n | n events |
| **MCI/Dementia** |  |  |  |  |
| Model 1 | 1.06 (1.00-1.13) | 0.06 | 6069 | 1307 |
| Model 2 | 1.04 (0.98-1.11) | 0.17 | 6069 | 1307 |
| Model 3 | 1.03 (0.97-1.10) | 0.32 | 6069 | 1307 |
| Model 4 | 1.02 (0.95-1.09) | 0.56 | 6069 | 1307 |
| **MCI** |  |  |  |  |
| Model 1 | 1.08 (1.00-1.16) | 0.07 | 6069 | 799 |
| Model 2 | 1.05 (0.98-1.14) | 0.18 | 6069 | 799 |
| Model 3 | 1.03 (0.95-1.12) | 0.45 | 6069 | 799 |
| Model 4 | 1.04 (0.95-1.13) | 0.38 | 6069 | 799 |
| **Dementia** |  |  |  |  |
| Model 1 | 1.04 (0.96-1.13) | 0.31 | 6069 | 786 |
| Model 2 | 1.03 (0.95-1.11) | 0.48 | 6069 | 786 |
| Model 3 | 1.02 (0.94-1.11) | 0.56 | 6069 | 786 |
| Model 4 | 0.99 (0.91-1.08) | 0.84 | 6069 | 786 |
|  | **DNAm GDF-15** |  |  |  |
|  | HR (95% CI) for 1-SD increment (97.2) | P-value^a^ | n | n events |
| **MCI/Dementia** |  |  |  |  |
| Model 1 | 1.00 (0.95-1.06) | 0.90 | 6069 | 1307 |
| Model 2 | 1.00 (0.95-1.06) | 0.91 | 6069 | 1307 |
| Model 3 | 0.99 (0.93-1.05) | 0.73 | 6069 | 1307 |
| Model 4 | 0.98 (0.92-1.05) | 0.58 | 6069 | 1307 |
| **MCI** |  |  |  |  |
| Model 1 | 0.99 (0.92-1.07) | 0.75 | 6069 | 799 |
| Model 2 | 0.99 (0.91-1.07) | 0.74 | 6069 | 799 |
| Model 3 | 0.97 (0.90-1.05) | 0.50 | 6069 | 799 |
| Model 4 | 0.98 (0.90-1.06) | 0.62 | 6069 | 799 |
| **Dementia** |  |  |  |  |
| Model 1 | 1.00 (0.92-1.08) | 0.94 | 6069 | 786 |
| Model 2 | 1.00 (0.92-1.08) | 0.94 | 6069 | 786 |
| Model 3 | 0.99 (0.91-1.07) | 0.79 | 6069 | 786 |
| Model 4 | 0.96 (0.89-1.05) | 0.40 | 6069 | 786 |
|  | **DNAm leptin** |  |  |  |
|  | HR (95% CI) for 1-SD increment (1,732) | P-value^a^ | n | n events |
| **MCI/Dementia** |  |  |  |  |
| Model 1 | 1.04 (0.99-1.10) | 0.15 | 6069 | 1307 |
| Model 2 | 1.04 (0.98-1.09) | 0.21 | 6069 | 1307 |
| Model 3 | 1.03 (0.97-1.09) | 0.35 | 6069 | 1307 |
| Model 4 | 1.02 (0.96-1.08) | 0.52 | 6069 | 1307 |
| **MCI** |  |  |  |  |
| Model 1 | 1.06 (0.99-1.14) | 0.08 | 6069 | 799 |
| Model 2 | 1.06 (0.98-1.13) | 0.13 | 6069 | 799 |
| Model 3 | 1.04 (0.97-1.12) | 0.27 | 6069 | 799 |
| Model 4 | 1.04 (0.96-1.12) | 0.32 | 6069 | 799 |
| **Dementia** |  |  |  |  |
| Model 1 | 1.05 (0.98-1.13) | 0.17 | 6069 | 786 |
| Model 2 | 1.05 (0.97-1.12) | 0.21 | 6069 | 786 |
| Model 3 | 1.04 (0.97-1.12) | 0.28 | 6069 | 786 |
| Model 4 | 1.03 (0.95-1.11) | 0.49 | 6069 | 786 |
|  | **DNAm log HbA1C** |  |  |  |
|  | HR (95% CI) for 1-SD increment (0.029) | P-value^a^ | n | n events |
| **MCI/Dementia** |  |  |  |  |
| Model 1 | 1.11 (1.04-1.17) | <0.001 | 6069 | 1307 |
| Model 2 | 1.07 (1.01-1.13) | 0.02 | 6069 | 1307 |
| Model 3 | 1.07 (1.01-1.13) | 0.03 | 6069 | 1307 |
| Model 4 | 1.07 (1.00-1.13) | 0.04 | 6069 | 1307 |
| **MCI** |  |  |  |  |
| Model 1 | 1.11 (1.04-1.20) | 0.004 | 6069 | 799 |
| Model 2 | 1.05 (0.98-1.13) | 0.17 | 6069 | 799 |
| Model 3 | 1.04 (0.97-1.13) | 0.27 | 6069 | 799 |
| Model 4 | 1.04 (0.96-1.12) | 0.35 | 6069 | 799 |
| **Dementia** |  |  |  |  |
| Model 1 | 1.07 (1.00-1.15) | 0.06 | 6069 | 786 |
| Model 2 | 1.06 (0.99-1.14) | 0.12 | 6069 | 786 |
| Model 3 | 1.06 (0.98-1.15) | 0.15 | 6069 | 786 |
| Model 4 | 1.06 (0.98-1.15) | 0.13 | 6069 | 786 |
|  | **DNAm CRP** |  |  |  |
|  | HR (95% CI) for 1-SD increment (0.40) | P-value^a^ | n | n events |
| **MCI/Dementia** |  |  |  |  |
| Model 1 | 1.10 (1.04-1.16) | 0.001 | 6069 | 1307 |
| Model 2 | 1.07 (1.01-1.13) | 0.02 | 6069 | 1307 |
| Model 3 | 1.07 (1.01-1.14) | 0.02 | 6069 | 1307 |
| Model 4 | 1.07 (1.00-1.14) | 0.06 | 6069 | 1307 |
| **MCI** |  |  |  |  |
| Model 1 | 1.11 (1.03-1.19) | 0.005 | 6069 | 799 |
| Model 2 | 1.06 (0.99-1.14) | 0.01 | 6069 | 799 |
| Model 3 | 1.06 (0.99-1.14) | 0.11 | 6069 | 799 |
| Model 4 | 1.08 (0.99-1.17) | 0.09 | 6069 | 799 |
| **Dementia** |  |  |  |  |
| Model 1 | 1.09 (1.02-1.17) | 0.02 | 6069 | 786 |
| Model 2 | 1.08 (1.00-1.16) | 0.04 | 6069 | 786 |
| Model 3 | 1.08 (1.00-1.16) | 0.05 | 6069 | 786 |
| Model 4 | 1.04 (0.95-1.14) | 0.37 | 6069 | 786 |
|  | **DNAm PACKYRS** |  |  |  |
|  | HR (95% CI) for 1-SD increment (10.7) | P-value^a^ | n | n events |
| **MCI/Dementia** |  |  |  |  |
| Model 1 | 1.09 (1.03-1.16) | 0.003 | 6069 | 1307 |
| Model 2 | 1.07 (1.01-1.14) | 0.03 | 6069 | 1307 |
| Model 3 | 1.09 (1.00-1.18) | 0.05 | 6069 | 1307 |
| Model 4 | 1.08 (0.99-1.17) | 0.07 | 6069 | 1307 |
| **MCI** |  |  |  |  |
| Model 1 | 1.08 (1.00-1.17) | 0.04 | 6069 | 799 |
| Model 2 | 1.04 (0.96-1.12) | 0.38 | 6069 | 799 |
| Model 3 | 1.06 (0.96-1.18) | 0.24 | 6069 | 799 |
| Model 4 | 1.07 (0.96-1.19) | 0.22 | 6069 | 799 |
| **Dementia** |  |  |  |  |
| Model 1 | 1.10 (1.02-1.19) | 0.01 | 6069 | 786 |
| Model 2 | 1.10 (1.02-1.19) | 0.02 | 6069 | 786 |
| Model 3 | 1.15 (1.03-1.28) | 0.01 | 6069 | 786 |
| Model 4 | 1.13 (1.01-1.26) | 0.03 | 6069 | 786 |
|  | **DNAm PAI-1** |  |  |  |
|  | HR (95% CI) for 1-SD increment (2,509) | P-value^a^ | n | n events |
| **MCI/Dementia** |  |  |  |  |
| Model 1 | 1.10 (1.04-1.17) | 0.001 | 6069 | 1307 |
| Model 2 | 1.09 (1.03-1.16) | 0.002 | 6069 | 1307 |
| Model 3 | 1.10 (1.04-1.16) | 0.002 | 6069 | 1307 |
| Model 4 | 1.10 (1.04-1.17) | 0.002 | 6069 | 1307 |
| **MCI** |  |  |  |  |
| Model 1 | 1.12 (1.04-1.20) | 0.002 | 6069 | 799 |
| Model 2 | 1.10 (1.03-1.18) | 0.008 | 6069 | 799 |
| Model 3 | 1.11 (1.03-1.19) | 0.008 | 6069 | 799 |
| Model 4 | 1.10 (1.02-1.19) | 0.01 | 6069 | 799 |
| **Dementia** |  |  |  |  |
| Model 1 | 1.08 (1.00-1.16) | 0.04 | 6069 | 786 |
| Model 2 | 1.08 (1.00-1.16) | 0.05 | 6069 | 786 |
| Model 3 | 1.08 (1.00-1.16) | 0.06 | 6069 | 786 |
| Model 4 | 1.08 (1.00-1.16) | 0.06 | 6069 | 786 |
|  | **DNAm TIMP-1** |  |  |  |
|  | HR (95% CI) for 1-SD increment (829) | P-value^a^ | n | n events |
| **MCI/Dementia** |  |  |  |  |
| Model 1 | 1.05 (0.98-1.12) | 0.14 | 6069 | 1307 |
| Model 2 | 1.03 (0.96-1.10) | 0.39 | 6069 | 1307 |
| Model 3 | 1.02 (0.95-1.09) | 0.59 | 6069 | 1307 |
| Model 4 | 1.00 (0.93-1.07) | 0.96 | 6069 | 1307 |
| **MCI** |  |  |  |  |
| Model 1 | 1.13 (1.04-1.23) | 0.004 | 6069 | 799 |
| Model 2 | 1.09 (1.01-1.18) | 0.04 | 6069 | 799 |
| Model 3 | 1.07 (0.99-1.17) | 0.010 | 6069 | 799 |
| Model 4 | 1.06 (0.97-1.16) | 0.22 | 6069 | 799 |
| **Dementia** |  |  |  |  |
| Model 1 | 0.99 (0.91-1.08) | 0.87 | 6069 | 786 |
| Model 2 | 0.98 (0.91-1.07) | 0.71 | 6069 | 786 |
| Model 3 | 0.98 (0.90-1.06) | 0.62 | 6069 | 786 |
| Model 4 | 0.95 (0.86-1.04) | 0.24 | 6069 | 786 |

Abbreviations: IEAA=intrinsic epigenetic age acceleration; EEAA=extrinsic epigenetic age acceleration
^a^P-values from Cox multivariate regression models with the epigenetic clock modeled in continuous form
Model 1 adjusted for age; model 2 adjusted for age, race and ethnicity; model 3 adjusted for model 2 covariates plus education, WHI hormone therapy trial arm, smoking, body mass index, and physical activity; model 4 adjusted for model 3 covariates plus estimated white blood cell (WBC) counts (CD8T, CD4T, natural killer cells, B cells, monocytes, and neutrophils). Models for IEAA, IEAA.Hannum, and EEAA did not contain white blood cell (WBC) counts because IEAA was derived to be independent of white blood cell counts while EEAA incorporates them into its calculation.

**eTable 5. Associations of epigenetic clocks with incident mild cognitive impairment (MCI)/probable dementia, MCI, and probable dementia stratified by age, race/ethnicity, hormone therapy regimen, menopause type, and APOE ε4 carriage in the Women’s Health Initiative Memory Study (n=6,069) 1995-2021.**

|  | AgeAccelHorvath | | | | | | | | | |
| --- | --- | --- | --- | --- | --- | --- | --- | --- | --- | --- |
|  |  | MCI/probable dementia | | | MCI | | | Probable dementia | | |
|  | n | n events | HR (95% CI) | P-int | n events | HR (95% CI) | P-int | n events | HR (95% CI) | P-int |
| Overall | 6069 | 1307 | 1.00 (0.94-1.06) |  | 799 | 1.04 (0.96-1.11) |  | 786 | 0.96 (0.89-1.04) |  |
| Age |  |  |  | 0.58 |  |  | 0.39 |  |  | 0.55 |
| > 69 years | 2470 | 456 | 1.02 (0.92-1.13) |  | 282 | 1.05 (0.93-1.2) |  | 253 | 1.02 (0.89-1.17) |  |
| ≥ 69 years | 3599 | 851 | 1.00 (0.93-1.07) |  | 517 | 1.03 (0.94-1.13) |  | 533 | 0.94 (0.86-1.03) |  |
| Race and ethnicity |  |  |  | 0.45 |  |  | 0.92 |  |  | 0.32 |
| Black | 414 | 130 | 1.00 (0.82-1.21) |  | 107 | 1.01 (0.81-1.25) |  | 56 | 1.14 (0.85-1.53) |  |
| Hispanic | 160 | 41 | 1.25 (0.79-1.99) |  | 24 | 1.05 (0.58-1.91) |  | 26 | 1.42 (0.76-2.66) |  |
| White | 5297 | 1096 | 0.98 (0.92-1.05) |  | 639 | 1.03 (0.95-1.12) |  | 686 | 0.94 (0.87-1.02) |  |
| E-alone trial |  |  |  | 0.57 |  |  | 0.08 |  |  | 0.33 |
| E-alone intervention | 1152 | 260 | 1.03 (0.90-1.17) |  | 181 | 0.92 (0.78-1.08) |  | 144 | 1.15 (0.97-1.38) |  |
| E-alone placebo | 1163 | 270 | 1.05 (0.93-1.19) |  | 169 | 1.10 (0.95-1.28) |  | 152 | 0.97 (0.82-1.14) |  |
| E + P trial |  |  |  | 0.97 |  |  | 0.82 |  |  | 0.61 |
| E + P intervention | 1854 | 404 | 0.98 (0.88-1.09) |  | 233 | 1.09 (0.95-1.26) |  | 260 | 0.87 (0.76-1.00) |  |
| E + P placebo | 1900 | 373 | 0.97 (0.87-1.08) |  | 216 | 1.02 (0.89-1.17) |  | 230 | 0.95 (0.83-1.09) |  |
| Menopause type |  |  |  | 0.88 |  |  | 0.95 |  |  | 0.93 |
| Natural | 4753 | 1007 | 1.01 (0.95-1.08) |  | 602 | 1.06 (0.98-1.15) |  | 621 | 0.98 (0.90-1.06) |  |
| Surgical | 451 | 93 | 1.09 (0.89-1.35) |  | 59 | 1.08 (0.83-1.42) |  | 49 | 1.12 (0.82-1.51) |  |
| *APOE ε4* carriage |  |  |  | 0.22 |  |  | 0.84 |  |  | 0.21 |
| 0 *ε4 alleles* | 3858 | 724 | 1.00 (0.93-1.08) |  | 440 | 1.02 (0.93-1.13) |  | 436 | 0.96 (0.87-1.06) |  |
| ≥ 1 *ε4 alleles* | 1306 | 357 | 0.93 (0.83-1.05) |  | 190 | 1.01 (0.87-1.19) |  | 238 | 0.88 (0.76-1.01) |  |
|  | AgeAccelHannum | | | | | | | | | |
|  |  | MCI/probable dementia | | | MCI | | | Probable dementia | | |
|  | n | n events | HR (95% CI) | P-int | n events | HR (95% CI) | P-int | n events | HR (95% CI) | P-int |
| Overall | 6069 | 1307 | 1.01 (0.95-1.07) |  | 799 | 1.03 (0.95-1.11) |  | 786 | 0.96 (0.88-1.03) |  |
| Age |  |  |  | 0.37 |  |  | 0.99 |  |  | 0.48 |
| > 69 years | 2470 | 456 | 1.08 (0.97-1.20) |  | 282 | 1.13 (0.98-1.29) |  | 253 | 1.01 (0.88-1.17) |  |
| ≥ 69 years | 3599 | 851 | 0.97 (0.90-1.05) |  | 517 | 0.98 (0.89-1.08) |  | 533 | 0.93 (0.84-1.02) |  |
| Race and ethnicity |  |  |  | 0.68 |  |  | 0.69 |  |  | 0.67 |
| Black | 414 | 130 | 0.92 (0.75-1.13) |  | 107 | 0.94 (0.75-1.17) |  | 56 | 0.94 (0.69-1.27) |  |
| Hispanic | 160 | 41 | 0.77 (0.48-1.22) |  | 24 | 0.95 (0.54-1.66) |  | 26 | 0.66 (0.34-1.28) |  |
| White | 5297 | 1096 | 1.01 (0.94-1.08) |  | 639 | 1.04 (0.95-1.13) |  | 686 | 0.96 (0.88-1.04) |  |
| E-alone trial |  |  |  | 0.37 |  |  | 0.28 |  |  | 0.91 |
| E-alone intervention | 1152 | 260 | 1.03 (0.90-1.18) |  | 181 | 0.95 (0.80-1.12) |  | 144 | 1.12 (0.94-1.35) |  |
| E-alone placebo | 1163 | 270 | 1.06 (0.93-1.21) |  | 169 | 1.04 (0.88-1.22) |  | 152 | 1.00 (0.84-1.19) |  |
| E + P trial |  |  |  | 0.70 |  |  | 0.48 |  |  | 0.94 |
| E + P intervention | 1854 | 404 | 0.99 (0.89-1.10) |  | 233 | 1.14 (0.99-1.31) |  | 260 | 0.87 (0.75-1.00) |  |
| E + P placebo | 1900 | 373 | 0.96 (0.85-1.08) |  | 216 | 0.95 (0.81-1.11) |  | 230 | 0.91 (0.78-1.07) |  |
| Menopause type |  |  |  | 0.58 |  |  | 0.68 |  |  | 0.23 |
| Natural | 4753 | 1007 | 1.03 (0.96-1.10) |  | 602 | 1.07 (0.98-1.17) |  | 621 | 0.96 (0.88-1.05) |  |
| Surgical | 451 | 93 | 1.26 (1.00-1.60) |  | 59 | 1.17 (0.86-1.60) |  | 49 | 1.31 (0.95-1.82) |  |
| *APOE ε4* carriage |  |  |  | 0.21 |  |  | 0.88 |  |  | 0.17 |
| 0 *ε4 alleles* | 3858 | 724 | 1.05 (0.96-1.14) |  | 440 | 1.05 (0.95-1.17) |  | 436 | 1.01 (0.91-1.12) |  |
| ≥ 1 *ε4 alleles* | 1306 | 357 | 0.93 (0.82-1.05) |  | 190 | 1.06 (0.90-1.25) |  | 238 | 0.84 (0.72-0.97) |  |
|  | IEAA | | | | | | | | | |
|  |  | MCI/probable dementia | | | MCI | | | Probable dementia | | |
|  | n | n events | HR (95% CI) | P-int | n events | HR (95% CI) | P-int | n events | HR (95% CI) | P-int |
| Overall | 6069 | 1307 | 0.98 (0.93-1.03) |  | 799 | 1.01 (0.94-1.08) |  | 786 | 0.96 (0.89-1.03) |  |
| Age |  |  |  | 0.66 |  |  | 0.81 |  |  | 0.12 |
| > 69 years | 2470 | 456 | 1.01 (0.92-1.12) |  | 282 | 1.04 (0.92-1.17) |  | 253 | 1.04 (0.91-1.18) |  |
| ≥ 69 years | 3599 | 851 | 0.97 (0.91-1.04) |  | 517 | 1.00 (0.91-1.09) |  | 533 | 0.93 (0.85-1.01) |  |
| Race and ethnicity |  |  |  | 0.92 |  |  | 0.95 |  |  | 0.74 |
| Black | 414 | 130 | 0.96 (0.80-1.14) |  | 107 | 0.97 (0.80-1.18) |  | 56 | 1.02 (0.77-1.36) |  |
| Hispanic | 160 | 41 | 1.06 (0.72-1.58) |  | 24 | 1.02 (0.61-1.70) |  | 26 | 1.08 (0.67-1.75) |  |
| White | 5297 | 1096 | 0.97 (0.92-1.03) |  | 639 | 1.01 (0.93-1.09) |  | 686 | 0.95 (0.88-1.02) |  |
| E-alone trial |  |  |  | 0.96 |  |  | 0.24 |  |  | 0.39 |
| E-alone intervention | 1152 | 260 | 1.02 (0.90-1.15) |  | 181 | 0.93 (0.80-1.08) |  | 144 | 1.12 (0.95-1.33) |  |
| E-alone placebo | 1163 | 270 | 1.03 (0.91-1.16) |  | 169 | 1.06 (0.91-1.23) |  | 152 | 1.00 (0.85-1.18) |  |
| E + P trial |  |  |  | 0.82 |  |  | 0.60 |  |  | 0.67 |
| E + P intervention | 1854 | 404 | 0.97 (0.88-1.07) |  | 233 | 1.05 (0.91-1.19) |  | 260 | 0.90 (0.80-1.03) |  |
| E + P placebo | 1900 | 373 | 0.93 (0.84-1.04) |  | 216 | 0.99 (0.86-1.13) |  | 230 | 0.91 (0.80-1.04) |  |
| Menopause type |  |  |  | >0.99 |  |  | 0.98 |  |  | 0.69 |
| Natural | 4753 | 1007 | 1.00 (0.94-1.06) |  | 602 | 1.04 (0.96-1.13) |  | 621 | 0.97 (0.90-1.05) |  |
| Surgical | 451 | 93 | 1.02 (0.82-1.27) |  | 59 | 1.05 (0.80-1.38) |  | 49 | 0.95 (0.69-1.30) |  |
| *APOE ε4* carriage |  |  |  | 0.48 |  |  | 0.69 |  |  | 0.72 |
| 0 *ε4 alleles* | 3858 | 724 | 0.98 (0.91-1.06) |  | 440 | 1.00 (0.91-1.10) |  | 436 | 0.95 (0.86-1.05) |  |
| ≥ 1 *ε4 alleles* | 1306 | 357 | 0.93 (0.84-1.04) |  | 190 | 0.97 (0.83-1.12) |  | 238 | 0.92 (0.81-1.05) |  |
|  | EEAA | | | | | | | | | |
|  |  | MCI/probable dementia | | | MCI | | | Probable dementia | | |
|  | n | n events | HR (95% CI) | P-int | n events | HR (95% CI) | P-int | n events | HR (95% CI) | P-int |
| Overall | 6069 | 1307 | 1.01 (0.96-1.07) |  | 799 | 1.02 (0.95-1.10) |  | 786 | 0.97 (0.91-1.05) |  |
| Age |  |  |  | 0.19 |  |  | 0.95 |  |  | 0.30 |
| > 69 years | 2470 | 456 | 1.02 (0.92-1.12) |  | 282 | 1.08 (0.95-1.23) |  | 253 | 0.95 (0.83-1.08) |  |
| ≥ 69 years | 3599 | 851 | 1.01 (0.94-1.08) |  | 517 | 1.00 (0.91-1.09) |  | 533 | 0.98 (0.90-1.07) |  |
| Race and ethnicity |  |  |  | 0.88 |  |  | 0.46 |  |  | 0.83 |
| Black | 414 | 130 | 0.95 (0.79-1.14) |  | 107 | 0.93 (0.76-1.14) |  | 56 | 0.99 (0.75-1.30) |  |
| Hispanic | 160 | 41 | 0.81 (0.56-1.18) |  | 24 | 0.97 (0.62-1.52) |  | 26 | 0.71 (0.43-1.16) |  |
| White | 5297 | 1096 | 1.01 (0.95-1.08) |  | 639 | 1.03 (0.95-1.12) |  | 686 | 0.97 (0.90-1.05) |  |
| E-alone trial |  |  |  | 0.30 |  |  | 0.32 |  |  | 0.81 |
| E-alone intervention | 1152 | 260 | 0.99 (0.88-1.12) |  | 181 | 0.92 (0.79-1.07) |  | 144 | 1.06 (0.91-1.25) |  |
| E-alone placebo | 1163 | 270 | 1.06 (0.94-1.21) |  | 169 | 1.03 (0.88-1.20) |  | 152 | 1.02 (0.87-1.21) |  |
| E + P trial |  |  |  | 0.81 |  |  | 0.94 |  |  | 0.69 |
| E + P intervention | 1854 | 404 | 1.01 (0.91-1.11) |  | 233 | 1.07 (0.94-1.21) |  | 260 | 0.95 (0.83-1.07) |  |
| E + P placebo | 1900 | 373 | 0.99 (0.89-1.11) |  | 216 | 1.05 (0.91-1.22) |  | 230 | 0.92 (0.80-1.05) |  |
| Menopause type |  |  |  | 0.51 |  |  | 0.68 |  |  | 0.21 |
| Natural | 4753 | 1007 | 1.03 (0.96-1.10) |  | 602 | 1.06 (0.98-1.16) |  | 621 | 0.98 (0.90-1.06) |  |
| Surgical | 451 | 93 | 1.24 (0.99-1.56) |  | 59 | 1.10 (0.82-1.47) |  | 49 | 1.34 (0.98-1.84) |  |
| *APOE ε4* carriage |  |  |  | 0.26 |  |  | 0.72 |  |  | 0.18 |
| 0 *ε4 alleles* | 3858 | 724 | 1.04 (0.97-1.13) |  | 440 | 1.03 (0.93-1.14) |  | 436 | 1.01 (0.91-1.12) |  |
| ≥ 1 *ε4 alleles* | 1306 | 357 | 0.97 (0.87-1.07) |  | 190 | 1.08 (0.94-1.24) |  | 238 | 0.89 (0.79-1.01) |  |
|  | IEAA.Hannum | | | | | | | | | |
|  |  | MCI/probable dementia | | | MCI | | | Probable dementia | | |
|  | n | n events | HR (95% CI) | P-int | n events | HR (95% CI) | P-int | n events | HR (95% CI) | P-int |
| Overall | 6069 | 1307 | 0.98 (0.92-1.03) |  | 799 | 0.99 (0.92-1.06) |  | 786 | 0.94 (0.88-1.02) |  |
| Age |  |  |  | 0.88 |  |  | 0.96 |  |  | 0.39 |
| > 69 years | 2470 | 456 | 1.07 (0.98-1.18) |  | 282 | 1.08 (0.96-1.22) |  | 253 | 1.05 (0.93-1.19) |  |
| ≥ 69 years | 3599 | 851 | 0.92 (0.86-0.99) |  | 517 | 0.94 (0.85-1.02) |  | 533 | 0.89 (0.81-0.97) |  |
| Race and ethnicity |  |  |  | 0.11 |  |  | 0.30 |  |  | 0.46 |
| Black | 414 | 130 | 0.86 (0.72-1.03) |  | 107 | 0.88 (0.72-1.07) |  | 56 | 0.82 (0.62-1.09) |  |
| Hispanic | 160 | 41 | 0.66 (0.44-1.00) |  | 24 | 0.78 (0.48-1.28) |  | 26 | 0.77 (0.43-1.38) |  |
| White | 5297 | 1096 | 0.99 (0.93-1.06) |  | 639 | 1.01 (0.93-1.09) |  | 686 | 0.96 (0.88-1.04) |  |
| E-alone trial |  |  |  | 0.73 |  |  | 0.77 |  |  | 0.80 |
| E-alone intervention | 1152 | 260 | 1.03 (0.91-1.17) |  | 181 | 0.98 (0.84-1.14) |  | 144 | 1.08 (0.91-1.28) |  |
| E-alone placebo | 1163 | 270 | 1.05 (0.93-1.18) |  | 169 | 1.00 (0.86-1.16) |  | 152 | 1.07 (0.91-1.26) |  |
| E + P trial |  |  |  | 0.54 |  |  | 0.07 |  |  | 0.98 |
| E + P intervention | 1854 | 404 | 0.96 (0.86-1.07) |  | 233 | 1.08 (0.94-1.23) |  | 260 | 0.87 (0.76-1.00) |  |
| E + P placebo | 1900 | 373 | 0.91 (0.81-1.02) |  | 216 | 0.89 (0.77-1.04) |  | 230 | 0.87 (0.75-1.00) |  |
| Menopause type |  |  |  | 0.33 |  |  | 0.62 |  |  | 0.41 |
| Natural | 4753 | 1007 | 0.99 (0.93-1.06) |  | 602 | 1.03 (0.94-1.12) |  | 621 | 0.94 (0.86-1.02) |  |
| Surgical | 451 | 93 | 1.17 (0.94-1.47) |  | 59 | 1.16 (0.88-1.54) |  | 49 | 1.16 (0.84-1.59) |  |
| *APOE ε4* carriage |  |  |  | 0.14 |  |  | 0.54 |  |  | 0.18 |
| 0 *ε4 alleles* | 3858 | 724 | 1.03 (0.95-1.11) |  | 440 | 1.04 (0.94-1.14) |  | 436 | 0.99 (0.89-1.09) |  |
| ≥ 1 *ε4 alleles* | 1306 | 357 | 0.92 (0.83-1.03) |  | 190 | 0.97 (0.84-1.13) |  | 238 | 0.87 (0.76-1.00) |  |
|  | AgeAccelPheno | | | | | | | | | |
|  |  | MCI/probable dementia | | | MCI | | | Probable dementia | | |
|  | n | n events | HR (95% CI) | P-int | n events | HR (95% CI) | P-int | n events | HR (95% CI) | P-int |
| Overall | 6069 | 1307 | 1.01 (0.95-1.07) |  | 799 | 0.99 (0.92-1.07) |  | 786 | 1.02 (0.94-1.10) |  |
| Age |  |  |  | 0.34 |  |  | 0.41 |  |  | 0.18 |
| > 69 years | 2470 | 456 | 1.07 (0.97-1.19) |  | 282 | 1.04 (0.92-1.19) |  | 253 | 1.12 (0.97-1.28) |  |
| ≥ 69 years | 3599 | 851 | 0.99 (0.91-1.06) |  | 517 | 0.97 (0.89-1.07) |  | 533 | 0.98 (0.89-1.08) |  |
| Race and ethnicity |  |  |  | 0.17 |  |  | 0.24 |  |  | 0.60 |
| Black | 414 | 130 | 0.91 (0.76-1.08) |  | 107 | 0.94 (0.78-1.14) |  | 56 | 0.89 (0.69-1.14) |  |
| Hispanic | 160 | 41 | 1.24 (0.82-1.87) |  | 24 | 1.31 (0.77-2.22) |  | 26 | 1.13 (0.66-1.93) |  |
| White | 5297 | 1096 | 1.03 (0.96-1.10) |  | 639 | 1.02 (0.93-1.11) |  | 686 | 1.03 (0.94-1.12) |  |
| E-alone trial |  |  |  | 0.02 |  |  | 0.01 |  |  | 0.65 |
| E-alone intervention | 1152 | 260 | 0.90 (0.79-1.03) |  | 181 | 0.80 (0.69-0.94) |  | 144 | 1.05 (0.89-1.25) |  |
| E-alone placebo | 1163 | 270 | 1.16 (1.02-1.33) |  | 169 | 1.14 (0.96-1.35) |  | 152 | 1.15 (0.95-1.38) |  |
| E + P trial |  |  |  | 0.39 |  |  | 0.69 |  |  | 0.69 |
| E + P intervention | 1854 | 404 | 1.06 (0.95-1.19) |  | 233 | 1.14 (0.98-1.32) |  | 260 | 1.00 (0.87-1.15) |  |
| E + P placebo | 1900 | 373 | 0.95 (0.85-1.06) |  | 216 | 0.95 (0.82-1.10) |  | 230 | 0.95 (0.82-1.10) |  |
| Menopause type |  |  |  | 0.89 |  |  | 0.56 |  |  | 0.25 |
| Natural | 4753 | 1007 | 1.00 (0.93-1.07) |  | 602 | 0.99 (0.91-1.08) |  | 621 | 1.01 (0.93-1.11) |  |
| Surgical | 451 | 93 | 1.16 (0.93-1.45) |  | 59 | 1.04 (0.78-1.39) |  | 49 | 1.47 (1.07-2.04) |  |
| *APOE ε4* carriage |  |  |  | 0.91 |  |  | 0.34 |  |  | 0.87 |
| 0 *ε4 alleles* | 3858 | 724 | 1.03 (0.95-1.12) |  | 440 | 0.99 (0.89-1.10) |  | 436 | 1.05 (0.94-1.16) |  |
| ≥ 1 *ε4 alleles* | 1306 | 357 | 1.02 (0.90-1.15) |  | 190 | 1.09 (0.92-1.30) |  | 238 | 0.99 (0.85-1.15) |  |
|  | AgeAccelGrim2 | | | | | | | | | |
|  |  | MCI/probable dementia | | | MCI | | | Probable dementia | | |
|  | n | n events | HR (95% CI) | P-int | n events | HR (95% CI) | P-int | n events | HR (95% CI) | P-int |
| Overall | 6069 | 1307 | 1.11 (1.02-1.20) |  | 799 | 1.1 (1.00-1.22) |  | 786 | 1.11 (1.00-1.23) |  |
| Age |  |  |  | 0.18 |  |  | 0.085 |  |  | 0.42 |
| > 69 years | 2470 | 456 | 1.07 (0.93-1.22) |  | 282 | 1.06 (0.89-1.26) |  | 253 | 1.06 (0.88-1.27) |  |
| ≥ 69 years | 3599 | 851 | 1.12 (1.02-1.24) |  | 517 | 1.11 (0.98-1.26) |  | 533 | 1.14 (1.00-1.29) |  |
| Race and ethnicity |  |  |  | 0.40 |  |  | 0.966 |  |  | 0.07 |
| Black | 414 | 130 | 1.11 (0.89-1.38) |  | 107 | 1.15 (0.90-1.47) |  | 56 | 0.87 (0.63-1.21) |  |
| Hispanic | 160 | 41 | 0.83 (0.43-1.62) |  | 24 | 1.18 (0.52-2.66) |  | 26 | 0.48 (0.15-1.53) |  |
| White | 5297 | 1096 | 1.12 (1.03-1.23) |  | 639 | 1.1 (0.98-1.23) |  | 686 | 1.16 (1.03-1.29) |  |
| E-alone trial |  |  |  | 0.15 |  |  | 0.64 |  |  | 0.17 |
| E-alone intervention | 1152 | 260 | 0.95 (0.80-1.14) |  | 181 | 1.04 (0.84-1.27) |  | 144 | 0.85 (0.66-1.09) |  |
| E-alone placebo | 1163 | 270 | 1.31 (1.11-1.55) |  | 169 | 1.34 (1.08-1.66) |  | 152 | 1.18 (0.94-1.49) |  |
| E + P trial |  |  |  | 0.11 |  |  | 0.003 |  |  | 0.76 |
| E + P intervention | 1854 | 404 | 1.07 (0.93-1.23) |  | 233 | 0.91 (0.76-1.10) |  | 260 | 1.25 (1.05-1.49) |  |
| E + P placebo | 1900 | 373 | 1.14 (0.98-1.32) |  | 216 | 1.21 (0.99-1.48) |  | 230 | 1.17 (0.96-1.42) |  |
| Menopause type |  |  |  | 0.78 |  |  | 0.87 |  |  | 0.73 |
| Natural | 4753 | 1007 | 1.11 (1.01-1.21) |  | 602 | 1.07 (0.96-1.21) |  | 621 | 1.14 (1.01-1.28) |  |
| Surgical | 451 | 93 | 1.01 (0.72-1.41) |  | 59 | 1.15 (0.77-1.72) |  | 49 | 0.79 (0.48-1.32) |  |
| *APOE ε4* carriage |  |  |  | 0.36 |  |  | 0.69 |  |  | 0.42 |
| 0 *ε4 alleles* | 3858 | 724 | 1.18 (1.06-1.32) |  | 440 | 1.09 (0.95-1.25) |  | 436 | 1.23 (1.07-1.42) |  |
| ≥ 1 *ε4 alleles* | 1306 | 357 | 1.03 (0.89-1.20) |  | 190 | 1.19 (0.97-1.47) |  | 238 | 1.00 (0.83-1.21) |  |
|  | DunedinPACE | | | | | | | | | |
|  |  | MCI/probable dementia | | | MCI | | | Probable dementia | | |
|  | n | n events | HR (95% CI) | P-int | n events | HR (95% CI) | P-int | n events | HR (95% CI) | P-int |
| Overall | 6026 | 1301 | 1.07 (1.01-1.15) |  | 797 | 1.11 (1.02-1.21) |  | 782 | 1.01 (0.93-1.10) |  |
| Age |  |  |  | 0.05 |  |  | 0.03 |  |  | 0.44 |
| > 69 years | 2452 | 454 | 1.06 (0.94-1.18) |  | 281 | 1.09 (0.95-1.26) |  | 252 | 0.97 (0.83-1.13) |  |
| ≥ 69 years | 3574 | 847 | 1.09 (1.00-1.19) |  | 516 | 1.12 (1.01-1.25) |  | 530 | 1.04 (0.93-1.15) |  |
| Race and ethnicity |  |  |  | 0.66 |  |  | 0.58 |  |  | 0.56 |
| Black | 410 | 129 | 0.99 (0.81-1.20) |  | 106 | 1.07 (0.87-1.32) |  | 56 | 0.81 (0.60-1.09) |  |
| Hispanic | 159 | 41 | 1.19 (0.77-1.83) |  | 24 | 1.69 (0.91-3.15) |  | 26 | 0.69 (0.39-1.22) |  |
| White | 5260 | 1091 | 1.09 (1.01-1.17) |  | 638 | 1.12 (1.01-1.23) |  | 682 | 1.03 (0.94-1.14) |  |
| E-alone trial |  |  |  | 0.95 |  |  | 0.81 |  |  | 0.47 |
| E-alone intervention | 1141 | 258 | 1.00 (0.85-1.16) |  | 179 | 1.05 (0.87-1.25) |  | 144 | 0.92 (0.75-1.14) |  |
| E-alone placebo | 1159 | 270 | 1.06 (0.92-1.22) |  | 169 | 1.15 (0.96-1.38) |  | 152 | 0.92 (0.76-1.11) |  |
| E + P trial |  |  |  | 0.54 |  |  | 0.07 |  |  | 0.06 |
| E + P intervention | 1835 | 401 | 1.16 (1.02-1.31) |  | 233 | 1.04 (0.88-1.22) |  | 257 | 1.24 (1.06-1.45) |  |
| E + P placebo | 1891 | 372 | 1.03 (0.91-1.17) |  | 216 | 1.17 (0.99-1.38) |  | 229 | 0.92 (0.78-1.09) |  |
| Menopause type |  |  |  | 0.63 |  |  | 0.28 |  |  | 0.50 |
| Natural | 4717 | 1002 | 1.09 (1.01-1.17) |  | 601 | 1.12 (1.01-1.23) |  | 617 | 1.04 (0.94-1.15) |  |
| Surgical | 448 | 93 | 1.22 (0.94-1.58) |  | 59 | 1.55 (1.13-2.13) |  | 49 | 0.80 (0.55-1.18) |  |
| *APOE ε4* carriage |  |  |  | 0.68 |  |  | 0.33 |  |  | 0.82 |
| 0 *ε4 alleles* | 3834 | 722 | 1.09 (0.99-1.20) |  | 440 | 1.08 (0.96-1.21) |  | 434 | 1.04 (0.92-1.18) |  |
| ≥ 1 *ε4 alleles* | 1292 | 354 | 1.11 (0.98-1.26) |  | 189 | 1.24 (1.04-1.48) |  | 236 | 1.03 (0.89-1.21) |  |
|  | PCHorvath1Resid | | | | | | | | | |
|  |  | MCI/probable dementia | | | MCI | | | Probable dementia | | |
|  | n | n events | HR (95% CI) | P-int | n events | HR (95% CI) | P-int | n events | HR (95% CI) | P-int |
| Overall | 6026 | 1301 | 0.97 (0.91-1.04) |  | 797 | 0.98 (0.90-1.07) |  | 782 | 0.98 (0.90-1.06) |  |
| Age |  |  |  | 0.77 |  |  | 0.83 |  |  | 0.16 |
| > 69 years | 2452 | 454 | 1.05 (0.94-1.18) |  | 281 | 1.04 (0.90-1.21) |  | 252 | 1.08 (0.92-1.25) |  |
| ≥ 69 years | 3574 | 847 | 0.95 (0.87-1.02) |  | 516 | 0.96 (0.87-1.06) |  | 530 | 0.94 (0.85-1.04) |  |
| Race and ethnicity |  |  |  | 0.89 |  |  | 0.67 |  |  | 0.92 |
| Black | 410 | 129 | 0.93 (0.75-1.15) |  | 106 | 0.97 (0.77-1.23) |  | 56 | 0.94 (0.68-1.32) |  |
| Hispanic | 159 | 41 | 1.00 (0.64-1.57) |  | 24 | 0.74 (0.40-1.36) |  | 26 | 1.28 (0.72-2.27) |  |
| White | 5260 | 1091 | 0.98 (0.91-1.05) |  | 638 | 0.98 (0.89-1.07) |  | 682 | 0.99 (0.90-1.08) |  |
| E-alone trial |  |  |  | 0.17 |  |  | 0.32 |  |  | 0.30 |
| E-alone intervention | 1141 | 258 | 0.98 (0.84-1.13) |  | 179 | 0.96 (0.80-1.14) |  | 144 | 1.00 (0.82-1.21) |  |
| E-alone placebo | 1159 | 270 | 1.04 (0.91-1.20) |  | 169 | 1.05 (0.89-1.25) |  | 152 | 1.01 (0.84-1.20) |  |
| E + P trial |  |  |  | 0.59 |  |  | 0.18 |  |  | 0.91 |
| E + P intervention | 1835 | 401 | 0.97 (0.86-1.10) |  | 233 | 0.96 (0.82-1.13) |  | 257 | 0.98 (0.84-1.13) |  |
| E + P placebo | 1891 | 372 | 0.92 (0.81-1.05) |  | 216 | 0.94 (0.79-1.11) |  | 229 | 0.96 (0.82-1.13) |  |
| Menopause type |  |  |  | 0.89 |  |  | 0.86 |  |  | 0.87 |
| Natural | 4717 | 1002 | 1.01 (0.94-1.09) |  | 601 | 0.99 (0.90-1.09) |  | 617 | 1.02 (0.93-1.12) |  |
| Surgical | 448 | 93 | 1.03 (0.81-1.31) |  | 59 | 1.01 (0.75-1.37) |  | 49 | 1.12 (0.80-1.57) |  |
| *APOE ε4* carriage |  |  |  | 0.11 |  |  | 0.96 |  |  | 0.38 |
| 0 *ε4 alleles* | 3834 | 722 | 1.02 (0.94-1.12) |  | 440 | 0.98 (0.87-1.10) |  | 434 | 1.03 (0.92-1.15) |  |
| ≥ 1 *ε4 alleles* | 1292 | 354 | 0.90 (0.80-1.02) |  | 189 | 0.99 (0.84-1.17) |  | 236 | 0.91 (0.78-1.05) |  |
|  | PCHannumResid | | | | | | | | | |
|  |  | MCI/probable dementia | | | MCI | | | Probable dementia | | |
|  | n | n events | HR (95% CI) | P-int | n events | HR (95% CI) | P-int | n events | HR (95% CI) | P-int |
| Overall | 6026 | 1301 | 0.98 (0.91-1.04) |  | 797 | 0.97 (0.89-1.06) |  | 782 | 0.98 (0.90-1.06) |  |
| Age |  |  |  | 0.51 |  |  | 0.51 |  |  | 0.56 |
| > 69 years | 2452 | 454 | 1.00 (0.89-1.12) |  | 281 | 0.98 (0.85-1.14) |  | 252 | 1.02 (0.87-1.19) |  |
| ≥ 69 years | 3574 | 847 | 0.97 (0.89-1.05) |  | 516 | 0.97 (0.88-1.08) |  | 530 | 0.96 (0.87-1.07) |  |
| Race and ethnicity |  |  |  | 0.92 |  |  | 0.69 |  |  | 0.95 |
| Black | 410 | 129 | 0.93 (0.75-1.15) |  | 106 | 0.99 (0.78-1.24) |  | 56 | 0.92 (0.66-1.28) |  |
| Hispanic | 159 | 41 | 0.96 (0.56-1.63) |  | 24 | 0.73 (0.37-1.43) |  | 26 | 1.13 (0.53-2.39) |  |
| White | 5260 | 1091 | 0.98 (0.91-1.05) |  | 638 | 0.97 (0.88-1.07) |  | 682 | 0.99 (0.90-1.09) |  |
| E-alone trial |  |  |  | 0.14 |  |  | 0.20 |  |  | 0.43 |
| E-alone intervention | 1141 | 258 | 0.95 (0.82-1.11) |  | 179 | 0.91 (0.76-1.09) |  | 144 | 1.00 (0.82-1.22) |  |
| E-alone placebo | 1159 | 270 | 1.05 (0.91-1.20) |  | 169 | 1.03 (0.86-1.23) |  | 152 | 1.02 (0.85-1.23) |  |
| E + P trial |  |  |  | 0.80 |  |  | 0.29 |  |  | 0.88 |
| E + P intervention | 1835 | 401 | 0.98 (0.87-1.11) |  | 233 | 1.00 (0.85-1.17) |  | 257 | 0.96 (0.82-1.12) |  |
| E + P placebo | 1891 | 372 | 0.94 (0.83-1.07) |  | 216 | 0.94 (0.80-1.12) |  | 229 | 0.97 (0.83-1.14) |  |
| Menopause type |  |  |  | 0.97 |  |  | 0.83 |  |  | 0.99 |
| Natural | 4717 | 1002 | 1.02 (0.94-1.10) |  | 601 | 0.99 (0.90-1.10) |  | 617 | 1.03 (0.93-1.13) |  |
| Surgical | 448 | 93 | 1.12 (0.88-1.41) |  | 59 | 1.1 (0.82-1.48) |  | 49 | 1.20 (0.85-1.70) |  |
| *APOE ε4* carriage |  |  |  | 0.11 |  |  | 0.89 |  |  | 0.47 |
| 0 *ε4 alleles* | 3834 | 722 | 1.03 (0.95-1.13) |  | 440 | 0.99 (0.88-1.11) |  | 434 | 1.04 (0.93-1.16) |  |
| ≥ 1 *ε4 alleles* | 1292 | 354 | 0.87 (0.77-1.00) |  | 189 | 0.95 (0.79-1.13) |  | 236 | 0.88 (0.75-1.03) |  |
|  | PCPhenoAgeResid | | | | | | | | | |
|  |  | MCI/probable dementia | | | MCI | | | Probable dementia | | |
|  | n | n events | HR (95% CI) | P-int | n events | HR (95% CI) | P-int | n events | HR (95% CI) | P-int |
| Overall | 6026 | 1301 | 1.05 (0.97-1.12) |  | 797 | 1.05 (0.95-1.15) |  | 782 | 1.02 (0.92-1.12) |  |
| Age |  |  |  | 0.91 |  |  | 0.44 |  |  | 0.89 |
| > 69 years | 2452 | 454 | 1.05 (0.92-1.20) |  | 281 | 1.01 (0.85-1.20) |  | 252 | 1.08 (0.90-1.29) |  |
| ≥ 69 years | 3574 | 847 | 1.04 (0.95-1.14) |  | 516 | 1.05 (0.94-1.18) |  | 530 | 1.00 (0.89-1.12) |  |
| Race and ethnicity |  |  |  | 0.67 |  |  | 0.51 |  |  | 0.83 |
| Black | 410 | 129 | 1.01 (0.81-1.26) |  | 106 | 1.09 (0.86-1.39) |  | 56 | 0.92 (0.65-1.29) |  |
| Hispanic | 159 | 41 | 0.81 (0.42-1.56) |  | 24 | 0.49 (0.21-1.12) |  | 26 | 1.32 (0.48-3.66) |  |
| White | 5260 | 1091 | 1.05 (0.97-1.14) |  | 638 | 1.04 (0.94-1.16) |  | 682 | 1.03 (0.93-1.14) |  |
| E-alone trial |  |  |  | 0.42 |  |  | 0.54 |  |  | 0.99 |
| E-alone intervention | 1141 | 258 | 0.98 (0.83-1.16) |  | 179 | 0.97 (0.80-1.19) |  | 144 | 1.00 (0.80-1.25) |  |
| E-alone placebo | 1159 | 270 | 1.11 (0.94-1.30) |  | 169 | 1.09 (0.89-1.33) |  | 152 | 1.02 (0.82-1.27) |  |
| E + P trial |  |  |  | 0.60 |  |  | 0.08 |  |  | 0.84 |
| E + P intervention | 1835 | 401 | 1.06 (0.93-1.21) |  | 233 | 1.06 (0.89-1.26) |  | 257 | 1.04 (0.88-1.23) |  |
| E + P placebo | 1891 | 372 | 1.03 (0.89-1.19) |  | 216 | 1.03 (0.85-1.25) |  | 229 | 1.02 (0.85-1.24) |  |
| Menopause type |  |  |  | 0.42 |  |  | 0.32 |  |  | 0.90 |
| Natural | 4717 | 1002 | 1.10 (1.01-1.20) |  | 601 | 1.09 (0.98-1.22) |  | 617 | 1.08 (0.97-1.21) |  |
| Surgical | 448 | 93 | 0.99 (0.74-1.33) |  | 59 | 0.95 (0.66-1.37) |  | 49 | 1.12 (0.73-1.70) |  |
| *APOE ε4* carriage |  |  |  | 0.22 |  |  | 0.72 |  |  | 0.64 |
| 0 *ε4 alleles* | 3834 | 722 | 1.13 (1.02-1.25) |  | 440 | 1.06 (0.93-1.20) |  | 434 | 1.11 (0.97-1.26) |  |
| ≥ 1 *ε4 alleles* | 1292 | 354 | 0.94 (0.82-1.08) |  | 189 | 1.06 (0.88-1.28) |  | 236 | 0.90 (0.76-1.07) |  |
|  | PCGrimAgeResid | | | | | | | | | |
|  |  | MCI/probable dementia | | | MCI | | | Probable dementia | | |
|  | n | n events | HR (95% CI) | P-int | n events | HR (95% CI) | P-int | n events | HR (95% CI) | P-int |
| Overall | 6026 | 1301 | 1.11 (1.02-1.20) |  | 797 | 1.13 (1.02-1.26) |  | 782 | 1.10 (0.99-1.23) |  |
| Age |  |  |  | 0.73 |  |  | 0.34 |  |  | 0.62 |
| > 69 years | 2452 | 454 | 1.11 (0.96-1.28) |  | 281 | 1.09 (0.91-1.31) |  | 252 | 1.14 (0.94-1.38) |  |
| ≥ 69 years | 3574 | 847 | 1.11 (1.01-1.23) |  | 516 | 1.15 (1.01-1.31) |  | 530 | 1.10 (0.96-1.25) |  |
| Race and ethnicity |  |  |  | 0.38 |  |  | 0.652 |  |  | 0.47 |
| Black | 410 | 129 | 1.00 (0.77-1.31) |  | 106 | 1.08 (0.80-1.46) |  | 56 | 0.83 (0.56-1.24) |  |
| Hispanic | 159 | 41 | 0.92 (0.50-1.68) |  | 24 | 0.99 (0.48-2.06) |  | 26 | 0.55 (0.19-1.57) |  |
| White | 5260 | 1091 | 1.14 (1.04-1.25) |  | 638 | 1.17 (1.04-1.31) |  | 682 | 1.13 (1.01-1.27) |  |
| E-alone trial |  |  |  | 0.04 |  |  | 0.28 |  |  | 0.09 |
| E-alone intervention | 1141 | 258 | 0.86 (0.71-1.06) |  | 179 | 0.96 (0.76-1.21) |  | 144 | 0.77 (0.58-1.02) |  |
| E-alone placebo | 1159 | 270 | 1.31 (1.09-1.57) |  | 169 | 1.38 (1.10-1.74) |  | 152 | 1.16 (0.90-1.48) |  |
| E + P trial |  |  |  | 0.23 |  |  | 0.01 |  |  | 0.96 |
| E + P intervention | 1835 | 401 | 1.15 (1.00-1.31) |  | 233 | 1.05 (0.87-1.27) |  | 257 | 1.26 (1.06-1.50) |  |
| E + P placebo | 1891 | 372 | 1.07 (0.90-1.26) |  | 216 | 1.17 (0.95-1.45) |  | 229 | 1.08 (0.88-1.33) |  |
| Menopause type |  |  |  | 0.81 |  |  | 0.54 |  |  | 0.94 |
| Natural | 4717 | 1002 | 1.13 (1.03-1.24) |  | 601 | 1.13 (1.00-1.28) |  | 617 | 1.15 (1.02-1.30) |  |
| Surgical | 448 | 93 | 1.05 (0.75-1.47) |  | 59 | 1.09 (0.72-1.67) |  | 49 | 0.98 (0.60-1.60) |  |
| *APOE ε4* carriage |  |  |  | 0.97 |  |  | 0.70 |  |  | 0.49 |
| 0 *ε4 alleles* | 3834 | 722 | 1.18 (1.06-1.32) |  | 440 | 1.19 (1.03-1.37) |  | 434 | 1.15 (0.99-1.33) |  |
| ≥ 1 *ε4 alleles* | 1292 | 354 | 1.11 (0.94-1.30) |  | 189 | 1.24 (0.99-1.54) |  | 236 | 1.12 (0.92-1.38) |  |
|  | DNAm ADM | | | | | | | | | |
|  |  | MCI/probable dementia | | | MCI | | | Probable dementia | | |
|  | n | n events | HR (95% CI) | P-int | n events | HR (95% CI) | P-int | n events | HR (95% CI) | P-int |
| Overall | 6069 | 1307 | 1.06 (0.99-1.14) |  | 799 | 1.07 (0.98-1.16) |  | 786 | 1.03 (0.95-1.13) |  |
| Age |  |  |  | 0.32 |  |  | 0.27 |  |  | 0.41 |
| > 69 years | 2470 | 456 | 0.99 (0.88-1.12) |  | 282 | 1.02 (0.87-1.18) |  | 253 | 0.93 (0.79-1.10) |  |
| ≥ 69 years | 3599 | 851 | 1.09 (1.01-1.19) |  | 517 | 1.08 (0.98-1.20) |  | 533 | 1.09 (0.98-1.21) |  |
| Race and ethnicity |  |  |  | 0.23 |  |  | 0.32 |  |  | 0.05 |
| Black | 414 | 130 | 1.02 (0.82-1.26) |  | 107 | 1.08 (0.86-1.36) |  | 56 | 0.81 (0.58-1.13) |  |
| Hispanic | 160 | 41 | 1.09 (0.71-1.67) |  | 24 | 2.02 (1.04-3.92) |  | 26 | 0.58 (0.32-1.05) |  |
| White | 5297 | 1096 | 1.09 (1.01-1.17) |  | 639 | 1.06 (0.96-1.17) |  | 686 | 1.08 (0.98-1.18) |  |
| E-alone trial |  |  |  | 0.10 |  |  | 0.86 |  |  | 0.02 |
| E-alone intervention | 1152 | 260 | 1.00 (0.86-1.16) |  | 181 | 1.04 (0.86-1.24) |  | 144 | 0.91 (0.74-1.11) |  |
| E-alone placebo | 1163 | 270 | 1.32 (1.14-1.53) |  | 169 | 1.24 (1.04-1.49) |  | 152 | 1.33 (1.09-1.63) |  |
| E + P trial |  |  |  | 0.08 |  |  | 0.05 |  |  | 0.35 |
| E + P intervention | 1854 | 404 | 0.99 (0.88-1.12) |  | 233 | 0.99 (0.84-1.16) |  | 260 | 1.01 (0.87-1.18) |  |
| E + P placebo | 1900 | 373 | 1.06 (0.94-1.21) |  | 216 | 1.09 (0.92-1.28) |  | 230 | 1.05 (0.89-1.23) |  |
| Menopause type |  |  |  | 0.31 |  |  | 0.67 |  |  | 0.80 |
| Natural | 4753 | 1007 | 1.06 (0.98-1.14) |  | 602 | 1.05 (0.95-1.16) |  | 621 | 1.04 (0.94-1.15) |  |
| Surgical | 451 | 93 | 1.01 (0.77-1.32) |  | 59 | 1.05 (0.75-1.47) |  | 49 | 1.01 (0.67-1.51) |  |
| *APOE ε4* carriage |  |  |  | 0.61 |  |  | 0.91 |  |  | 0.90 |
| 0 *ε4 alleles* | 3858 | 724 | 1.11 (1.02-1.22) |  | 440 | 1.09 (0.97-1.23) |  | 436 | 1.09 (0.97-1.23) |  |
| ≥ 1 *ε4 alleles* | 1306 | 357 | 1.02 (0.90-1.17) |  | 190 | 1.06 (0.89-1.26) |  | 238 | 1.00 (0.85-1.17) |  |
|  | DNAm B2M | | | | | | | | | |
|  |  | MCI/probable dementia | | | MCI | | | Probable dementia | | |
|  | n | n events | HR (95% CI) | P-int | n events | HR (95% CI) | P-int | n events | HR (95% CI) | P-int |
| Overall | 6069 | 1307 | 1.01 (0.93-1.10) |  | 799 | 1.03 (0.93-1.15) |  | 786 | 0.94 (0.85-1.05) |  |
| Age |  |  |  | 0.23 |  |  | 0.32 |  |  | 0.17 |
| > 69 years | 2470 | 456 | 1.11 (0.95-1.29) |  | 282 | 1.01 (0.83-1.23) |  | 253 | 1.11 (0.91-1.37) |  |
| ≥ 69 years | 3599 | 851 | 0.97 (0.88-1.07) |  | 517 | 1.04 (0.92-1.18) |  | 533 | 0.89 (0.78-1.01) |  |
| Race and ethnicity |  |  |  | 0.66 |  |  | 0.72 |  |  | 0.43 |
| Black | 414 | 130 | 1.04 (0.82-1.31) |  | 107 | 1.08 (0.83-1.39) |  | 56 | 0.86 (0.60-1.22) |  |
| Hispanic | 160 | 41 | 0.69 (0.38-1.26) |  | 24 | 0.92 (0.46-1.84) |  | 26 | 0.29 (0.11-0.75) |  |
| White | 5297 | 1096 | 1.01 (0.92-1.10) |  | 639 | 1.00 (0.88-1.13) |  | 686 | 0.97 (0.87-1.09) |  |
| E-alone trial |  |  |  | 0.96 |  |  | 0.70 |  |  | 0.92 |
| E-alone intervention | 1152 | 260 | 0.95 (0.79-1.14) |  | 181 | 0.97 (0.78-1.21) |  | 144 | 0.92 (0.71-1.17) |  |
| E-alone placebo | 1163 | 270 | 1.17 (0.98-1.40) |  | 169 | 1.09 (0.87-1.36) |  | 152 | 1.07 (0.84-1.36) |  |
| E + P trial |  |  |  | 0.42 |  |  | 0.37 |  |  | 0.85 |
| E + P intervention | 1854 | 404 | 0.91 (0.78-1.05) |  | 233 | 0.98 (0.81-1.20) |  | 260 | 0.89 (0.74-1.08) |  |
| E + P placebo | 1900 | 373 | 1.05 (0.90-1.23) |  | 216 | 1.03 (0.84-1.27) |  | 230 | 0.99 (0.81-1.22) |  |
| Menopause type |  |  |  | 0.07 |  |  | 0.40 |  |  | 0.31 |
| Natural | 4753 | 1007 | 1.03 (0.94-1.13) |  | 602 | 1.07 (0.94-1.20) |  | 621 | 0.97 (0.86-1.09) |  |
| Surgical | 451 | 93 | 0.94 (0.69-1.29) |  | 59 | 0.99 (0.69-1.43) |  | 49 | 0.76 (0.45-1.30) |  |
| *APOE ε4* carriage |  |  |  | 0.28 |  |  | 0.39 |  |  | 0.38 |
| 0 *ε4 alleles* | 3858 | 724 | 1.09 (0.98-1.22) |  | 440 | 0.99 (0.86-1.15) |  | 436 | 1.11 (0.96-1.29) |  |
| ≥ 1 *ε4 alleles* | 1306 | 357 | 0.90 (0.76-1.05) |  | 190 | 1.10 (0.89-1.36) |  | 238 | 0.79 (0.64-0.97) |  |
|  | DNAm Cystatin C | | | | | | | | | |
|  |  | MCI/probable dementia | | | MCI | | | Probable dementia | | |
|  | n | n events | HR (95% CI) | P-int | n events | HR (95% CI) | P-int | n events | HR (95% CI) | P-int |
| Overall | 6069 | 1307 | 1.02 (0.95-1.09) |  | 799 | 1.04 (0.95-1.13) |  | 786 | 0.99 (0.91-1.08) |  |
| Age |  |  |  | 0.29 |  |  | 0.11 |  |  | 0.40 |
| > 69 years | 2470 | 456 | 1.04 (0.92-1.17) |  | 282 | 1.08 (0.92-1.25) |  | 253 | 1.00 (0.84-1.18) |  |
| ≥ 69 years | 3599 | 851 | 1.01 (0.93-1.09) |  | 517 | 1.02 (0.92-1.13) |  | 533 | 0.99 (0.89-1.10) |  |
| Race and ethnicity |  |  |  | 0.29 |  |  | 0.31 |  |  | 0.43 |
| Black | 414 | 130 | 0.96 (0.77-1.18) |  | 107 | 0.96 (0.76-1.22) |  | 56 | 1.00 (0.72-1.40) |  |
| Hispanic | 160 | 41 | 0.61 (0.34-1.08) |  | 24 | 0.59 (0.30-1.17) |  | 26 | 0.63 (0.25-1.63) |  |
| White | 5297 | 1096 | 1.03 (0.95-1.10) |  | 639 | 1.03 (0.94-1.14) |  | 686 | 1.01 (0.92-1.11) |  |
| E-alone trial |  |  |  | 0.51 |  |  | 0.81 |  |  | 0.48 |
| E-alone intervention | 1152 | 260 | 0.98 (0.84-1.13) |  | 181 | 0.98 (0.82-1.17) |  | 144 | 0.92 (0.75-1.12) |  |
| E-alone placebo | 1163 | 270 | 1.15 (0.99-1.33) |  | 169 | 1.12 (0.92-1.36) |  | 152 | 1.10 (0.90-1.34) |  |
| E + P trial |  |  |  | 0.95 |  |  | 0.65 |  |  | 0.53 |
| E + P intervention | 1854 | 404 | 1.01 (0.90-1.14) |  | 233 | 1.07 (0.92-1.25) |  | 260 | 1.03 (0.88-1.20) |  |
| E + P placebo | 1900 | 373 | 0.97 (0.86-1.11) |  | 216 | 0.99 (0.83-1.17) |  | 230 | 0.97 (0.82-1.15) |  |
| Menopause type |  |  |  | 0.58 |  |  | 0.67 |  |  | 0.53 |
| Natural | 4753 | 1007 | 1.04 (0.96-1.12) |  | 602 | 1.06 (0.96-1.16) |  | 621 | 1.01 (0.91-1.11) |  |
| Surgical | 451 | 93 | 1.09 (0.85-1.41) |  | 59 | 0.99 (0.72-1.38) |  | 49 | 1.40 (0.99-1.99) |  |
| *APOE ε4* carriage |  |  |  | 0.31 |  |  | 0.21 |  |  | 0.42 |
| 0 *ε4 alleles* | 3858 | 724 | 1.07 (0.98-1.17) |  | 440 | 1.00 (0.89-1.13) |  | 436 | 1.08 (0.96-1.21) |  |
| ≥ 1 *ε4 alleles* | 1306 | 357 | 0.97 (0.85-1.11) |  | 190 | 1.15 (0.97-1.36) |  | 238 | 0.91 (0.78-1.07) |  |
|  | DNAm GDF-15 | | | | | | | | | |
|  |  | MCI/probable dementia | | | MCI | | | Probable dementia | | |
|  | n | n events | HR (95% CI) | P-int | n events | HR (95% CI) | P-int | n events | HR (95% CI) | P-int |
| Overall | 6069 | 1307 | 0.98 (0.92-1.05) |  | 799 | 0.98 (0.90-1.06) |  | 786 | 0.96 (0.89-1.05) |  |
| Age |  |  |  | 0.30 |  |  | 0.52 |  |  | 0.73 |
| > 69 years | 2470 | 456 | 0.89 (0.79-1.02) |  | 282 | 0.88 (0.75-1.04) |  | 253 | 0.89 (0.75-1.06) |  |
| ≥ 69 years | 3599 | 851 | 1.02 (0.94-1.09) |  | 517 | 1.01 (0.92-1.11) |  | 533 | 1.00 (0.91-1.10) |  |
| Race and ethnicity |  |  |  | 0.28 |  |  | 0.18 |  |  | 0.83 |
| Black | 414 | 130 | 0.92 (0.74-1.14) |  | 107 | 0.88 (0.69-1.12) |  | 56 | 0.91 (0.66-1.25) |  |
| Hispanic | 160 | 41 | 0.53 (0.30-0.93) |  | 24 | 0.60 (0.29-1.24) |  | 26 | 0.43 (0.18-1.05) |  |
| White | 5297 | 1096 | 1.00 (0.94-1.08) |  | 639 | 1.00 (0.92-1.10) |  | 686 | 0.99 (0.90-1.08) |  |
| E-alone trial |  |  |  | 0.94 |  |  | 0.93 |  |  | 0.93 |
| E-alone intervention | 1152 | 260 | 0.92 (0.79-1.08) |  | 181 | 0.90 (0.74-1.09) |  | 144 | 0.92 (0.74-1.15) |  |
| E-alone placebo | 1163 | 270 | 0.96 (0.85-1.09) |  | 169 | 0.99 (0.84-1.15) |  | 152 | 0.91 (0.76-1.09) |  |
| E + P trial |  |  |  | 0.54 |  |  | 0.23 |  |  | 0.81 |
| E + P intervention | 1854 | 404 | 1.01 (0.90-1.14) |  | 233 | 1.00 (0.85-1.17) |  | 260 | 1.02 (0.88-1.19) |  |
| E + P placebo | 1900 | 373 | 1.02 (0.90-1.16) |  | 216 | 1.03 (0.87-1.22) |  | 230 | 0.99 (0.84-1.16) |  |
| Menopause type |  |  |  | 0.45 |  |  | 0.53 |  |  | 0.77 |
| Natural | 4753 | 1007 | 1.02 (0.95-1.10) |  | 602 | 1.01 (0.92-1.12) |  | 621 | 1.00 (0.91-1.11) |  |
| Surgical | 451 | 93 | 1.03 (0.78-1.35) |  | 59 | 0.88 (0.61-1.29) |  | 49 | 1.29 (0.93-1.79) |  |
| *APOE ε4* carriage |  |  |  | 0.44 |  |  | 0.35 |  |  | 0.74 |
| 0 *ε4 alleles* | 3858 | 724 | 1.03 (0.95-1.12) |  | 440 | 1.00 (0.90-1.11) |  | 436 | 1.02 (0.92-1.14) |  |
| ≥ 1 *ε4 alleles* | 1306 | 357 | 0.97 (0.84-1.11) |  | 190 | 1.10 (0.92-1.30) |  | 238 | 0.93 (0.79-1.11) |  |
|  | DNAm leptin | | | | | | | | | |
|  |  | MCI/probable dementia | | | MCI | | | Probable dementia | | |
|  | n | n events | HR (95% CI) | P-int | n events | HR (95% CI) | P-int | n events | HR (95% CI) | P-int |
| Overall | 6069 | 1307 | 1.02 (0.96-1.08) |  | 799 | 1.04 (0.96-1.12) |  | 786 | 1.03 (0.95-1.11) |  |
| Age |  |  |  | 0.29 |  |  | 0.87 |  |  | 0.13 |
| > 69 years | 2470 | 456 | 0.98 (0.88-1.08) |  | 282 | 0.95 (0.84-1.08) |  | 253 | 1.02 (0.89-1.16) |  |
| ≥ 69 years | 3599 | 851 | 1.04 (0.97-1.12) |  | 517 | 1.09 (1-1.2) |  | 533 | 1.03 (0.94-1.13) |  |
| Race and ethnicity |  |  |  | 0.91 |  |  | 0.29 |  |  | 0.93 |
| Black | 414 | 130 | 1.08 (0.89-1.31) |  | 107 | 1.23 (1.00-1.52) |  | 56 | 0.98 (0.73-1.30) |  |
| Hispanic | 160 | 41 | 1.10 (0.73-1.65) |  | 24 | 1.17 (0.70-1.96) |  | 26 | 0.87 (0.48-1.59) |  |
| White | 5297 | 1096 | 1.02 (0.96-1.09) |  | 639 | 1.01 (0.93-1.10) |  | 686 | 1.04 (0.96-1.13) |  |
| E-alone trial |  |  |  | 1.0 |  |  | 0.79 |  |  | 0.78 |
| E-alone intervention | 1152 | 260 | 1.07 (0.94-1.22) |  | 181 | 1.11 (0.94-1.30) |  | 144 | 1.11 (0.93-1.31) |  |
| E-alone placebo | 1163 | 270 | 1.11 (0.98-1.26) |  | 169 | 1.12 (0.95-1.32) |  | 152 | 1.15 (0.97-1.36) |  |
| E + P trial |  |  |  | 0.61 |  |  | 0.82 |  |  | 0.59 |
| E + P intervention | 1854 | 404 | 0.98 (0.88-1.09) |  | 233 | 0.98 (0.85-1.13) |  | 260 | 0.98 (0.86-1.13) |  |
| E + P placebo | 1900 | 373 | 0.97 (0.87-1.09) |  | 216 | 0.97 (0.84-1.12) |  | 230 | 0.98 (0.85-1.13) |  |
| Menopause type |  |  |  | 0.50 |  |  | 0.75 |  |  | 0.60 |
| Natural | 4753 | 1007 | 1.02 (0.96-1.10) |  | 602 | 1.03 (0.94-1.12) |  | 621 | 1.03 (0.95-1.12) |  |
| Surgical | 451 | 93 | 0.92 (0.75-1.13) |  | 59 | 1.04 (0.80-1.35) |  | 49 | 0.89 (0.67-1.20) |  |
| *APOE ε4* carriage |  |  |  | 0.34 |  |  | 0.61 |  |  | 0.33 |
| 0 *ε4 alleles* | 3858 | 724 | 1.02 (0.94-1.11) |  | 440 | 1.02 (0.92-1.12) |  | 436 | 1.03 (0.93-1.14) |  |
| ≥ 1 *ε4 alleles* | 1306 | 357 | 0.98 (0.87-1.09) |  | 190 | 1.01 (0.86-1.18) |  | 238 | 0.98 (0.85-1.13) |  |
|  | DNAm PACKYRS | | | | | | | | | |
|  |  | MCI/probable dementia | | | MCI | | | Probable dementia | | |
|  | n | n events | HR (95% CI) | P-int | n events | HR (95% CI) | P-int | n events | HR (95% CI) | P-int |
| Overall | 6069 | 1307 | 1.08 (0.99-1.17) |  | 799 | 1.07 (0.96-1.19) |  | 786 | 1.13 (1.01-1.26) |  |
| Age |  |  |  | 0.78 |  |  | 0.95 |  |  | 0.85 |
| > 69 years | 2470 | 456 | 1.07 (0.94-1.22) |  | 282 | 1.02 (0.86-1.22) |  | 253 | 1.15 (0.96-1.37) |  |
| ≥ 69 years | 3599 | 851 | 1.08 (0.97-1.20) |  | 517 | 1.09 (0.95-1.24) |  | 533 | 1.12 (0.97-1.28) |  |
| Race and ethnicity |  |  |  | 0.87 |  |  | 0.96 |  |  | 0.78 |
| Black | 414 | 130 | 1.20 (0.93-1.55) |  | 107 | 1.16 (0.87-1.55) |  | 56 | 1.11 (0.77-1.62) |  |
| Hispanic | 160 | 41 | 0.92 (0.50-1.71) |  | 24 | 1.04 (0.47-2.29) |  | 26 | 1.07 (0.41-2.81) |  |
| White | 5297 | 1096 | 1.07 (0.98-1.18) |  | 639 | 1.07 (0.95-1.20) |  | 686 | 1.12 (1.00-1.26) |  |
| E-alone trial |  |  |  | 0.39 |  |  | 0.71 |  |  | 0.51 |
| E-alone intervention | 1152 | 260 | 0.91 (0.74-1.12) |  | 181 | 0.94 (0.74-1.2) |  | 144 | 0.96 (0.71-1.29) |  |
| E-alone placebo | 1163 | 270 | 1.14 (0.97-1.35) |  | 169 | 1.23 (1.00-1.52) |  | 152 | 1.05 (0.83-1.33) |  |
| E + P trial |  |  |  | 0.55 |  |  | 0.10 |  |  | 0.84 |
| E + P intervention | 1854 | 404 | 1.09 (0.95-1.26) |  | 233 | 0.92 (0.76-1.12) |  | 260 | 1.27 (1.07-1.52) |  |
| E + P placebo | 1900 | 373 | 1.08 (0.92-1.28) |  | 216 | 1.16 (0.93-1.44) |  | 230 | 1.14 (0.92-1.42) |  |
| Menopause type |  |  |  | 0.95 |  |  | 0.93 |  |  | 0.84 |
| Natural | 4753 | 1007 | 1.07 (0.98-1.18) |  | 602 | 1.04 (0.92-1.18) |  | 621 | 1.15 (1.01-1.30) |  |
| Surgical | 451 | 93 | 1.00 (0.70-1.43) |  | 59 | 1.06 (0.67-1.67) |  | 49 | 0.78 (0.46-1.32) |  |
| *APOE ε4* carriage |  |  |  | 0.61 |  |  | 0.90 |  |  | 0.77 |
| 0 *ε4 alleles* | 3858 | 724 | 1.11 (0.99-1.24) |  | 440 | 1.08 (0.94-1.25) |  | 436 | 1.14 (0.98-1.32) |  |
| ≥ 1 *ε4 alleles* | 1306 | 357 | 1.02 (0.87-1.20) |  | 190 | 1.07 (0.86-1.34) |  | 238 | 1.09 (0.89-1.34) |  |
|  | DNAm PAI-1 | | | | | | | | | |
|  |  | MCI/probable dementia | | | MCI | | | Probable dementia | | |
|  | n | n events | HR (95% CI) | P-int | n events | HR (95% CI) | P-int | n events | HR (95% CI) | P-int |
| Overall | 6069 | 1307 | 1.10 (1.04-1.17) |  | 799 | 1.10 (1.02-1.19) |  | 786 | 1.08 (1.00-1.16) |  |
| Age |  |  |  | 0.46 |  |  | 0.29 |  |  | 0.59 |
| > 69 years | 2470 | 456 | 1.08 (0.98-1.20) |  | 282 | 1.08 (0.95-1.22) |  | 253 | 1.05 (0.92-1.21) |  |
| ≥ 69 years | 3599 | 851 | 1.11 (1.03-1.19) |  | 517 | 1.11 (1.01-1.22) |  | 533 | 1.09 (0.99-1.20) |  |
| Race and ethnicity |  |  |  | 0.94 |  |  | 0.48 |  |  | 0.61 |
| Black | 414 | 130 | 1.10 (0.91-1.32) |  | 107 | 1.15 (0.94-1.41) |  | 56 | 0.98 (0.73-1.32) |  |
| Hispanic | 160 | 41 | 1.45 (0.96-2.18) |  | 24 | 1.57 (0.91-2.69) |  | 26 | 1.14 (0.63-2.05) |  |
| White | 5297 | 1096 | 1.09 (1.03-1.17) |  | 639 | 1.08 (0.99-1.18) |  | 686 | 1.09 (1.00-1.18) |  |
| E-alone trial |  |  |  | 0.23 |  |  | 0.54 |  |  | 0.36 |
| E-alone intervention | 1152 | 260 | 1.08 (0.94-1.24) |  | 181 | 1.14 (0.97-1.34) |  | 144 | 1.03 (0.85-1.25) |  |
| E-alone placebo | 1163 | 270 | 1.30 (1.13-1.48) |  | 169 | 1.35 (1.14-1.60) |  | 152 | 1.22 (1.02-1.46) |  |
| E + P trial |  |  |  | 0.81 |  |  | 0.11 |  |  | 0.74 |
| E + P intervention | 1854 | 404 | 1.05 (0.94-1.17) |  | 233 | 0.94 (0.82-1.08) |  | 260 | 1.11 (0.97-1.26) |  |
| E + P placebo | 1900 | 373 | 1.05 (0.94-1.17) |  | 216 | 1.09 (0.95-1.26) |  | 230 | 1.03 (0.89-1.19) |  |
| Menopause type |  |  |  | 0.07 |  |  | 0.01 |  |  | 0.83 |
| Natural | 4753 | 1007 | 1.06 (0.99-1.13) |  | 602 | 1.02 (0.94-1.12) |  | 621 | 1.07 (0.98-1.17) |  |
| Surgical | 451 | 93 | 1.29 (1.02-1.63) |  | 59 | 1.63 (1.21-2.20) |  | 49 | 0.86 (0.6-1.23) |  |
| *APOE ε4* carriage |  |  |  | 0.14 |  |  | 0.49 |  |  | 0.08 |
| 0 *ε4 alleles* | 3858 | 724 | 1.12 (1.04-1.21) |  | 440 | 1.10 (0.99-1.22) |  | 436 | 1.13 (1.02-1.25) |  |
| ≥ 1 *ε4 alleles* | 1306 | 357 | 1.02 (0.91-1.14) |  | 190 | 1.05 (0.90-1.23) |  | 238 | 0.98 (0.85-1.13) |  |
|  | DNAm TIMP-1 | | | | | | | | | |
|  |  | MCI/probable dementia | | | MCI | | | Probable dementia | | |
|  | n | n events | HR (95% CI) | P-int | n events | HR (95% CI) | P-int | n events | HR (95% CI) | P-int |
| Overall | 6069 | 1307 | 1.00 (0.93-1.07) |  | 799 | 1.06 (0.97-1.16) |  | 786 | 0.95 (0.86-1.04) |  |
| Age |  |  |  | 0.10 |  |  | 0.02 |  |  | 0.20 |
| > 69 years | 2470 | 456 | 1.02 (0.9-1.16) |  | 282 | 1.12 (0.96-1.32) |  | 253 | 0.95 (0.80-1.13) |  |
| ≥ 69 years | 3599 | 851 | 0.98 (0.9-1.07) |  | 517 | 1.02 (0.91-1.14) |  | 533 | 0.94 (0.84-1.05) |  |
| Race and ethnicity |  |  |  | 0.44 |  |  | 0.42 |  |  | 0.56 |
| Black | 414 | 130 | 0.94 (0.76-1.16) |  | 107 | 1.04 (0.82-1.31) |  | 56 | 0.78 (0.56-1.09) |  |
| Hispanic | 160 | 41 | 1.05 (0.63-1.76) |  | 24 | 1.31 (0.70-2.47) |  | 26 | 0.75 (0.35-1.60) |  |
| White | 5297 | 1096 | 0.99 (0.92-1.08) |  | 639 | 1.04 (0.94-1.15) |  | 686 | 0.97 (0.87-1.07) |  |
| E-alone trial |  |  |  | 0.62 |  |  | 0.69 |  |  | 0.98 |
| E-alone intervention | 1152 | 260 | 0.99 (0.85-1.16) |  | 181 | 0.99 (0.82-1.19) |  | 144 | 0.91 (0.74-1.12) |  |
| E-alone placebo | 1163 | 270 | 1.09 (0.94-1.28) |  | 169 | 1.12 (0.92-1.36) |  | 152 | 1.03 (0.84-1.27) |  |
| E + P trial |  |  |  | 0.69 |  |  | 0.87 |  |  | 0.60 |
| E + P intervention | 1854 | 404 | 0.99 (0.87-1.14) |  | 233 | 1.12 (0.94-1.33) |  | 260 | 0.96 (0.81-1.13) |  |
| E + P placebo | 1900 | 373 | 0.93 (0.81-1.07) |  | 216 | 1.02 (0.85-1.22) |  | 230 | 0.90 (0.75-1.07) |  |
| Menopause type |  |  |  | 0.83 |  |  | 0.98 |  |  | 0.48 |
| Natural | 4753 | 1007 | 1.03 (0.95-1.12) |  | 602 | 1.11 (1.00-1.23) |  | 621 | 0.98 (0.88-1.09) |  |
| Surgical | 451 | 93 | 1.15 (0.87-1.52) |  | 59 | 1.10 (0.78-1.56) |  | 49 | 1.39 (0.94-2.04) |  |
| *APOE ε4* carriage |  |  |  | 0.68 |  |  | 0.38 |  |  | 0.74 |
| 0 *ε4 alleles* | 3858 | 724 | 1.02 (0.92-1.13) |  | 440 | 1.02 (0.89-1.16) |  | 436 | 0.97 (0.85-1.11) |  |
| ≥ 1 *ε4 alleles* | 1306 | 357 | 0.95 (0.83-1.09) |  | 190 | 1.11 (0.93-1.34) |  | 238 | 0.93 (0.79-1.09) |  |
|  | DNAm logA1C | | | | | | | | | |
|  |  | MCI/probable dementia | | | MCI | | | Probable dementia | | |
|  | n | n events | HR (95% CI) | P-int | n events | HR (95% CI) | P-int | n events | HR (95% CI) | P-int |
| Overall | 6069 | 1307 | 1.07 (1.00-1.13) |  | 799 | 1.04 (0.96-1.12) |  | 786 | 1.06 (0.98-1.15) |  |
| Age |  |  |  | 0.22 |  |  | 0.06 |  |  | 0.23 |
| > 69 years | 2470 | 456 | 1.08 (0.97-1.19) |  | 282 | 1.07 (0.94-1.21) |  | 253 | 1.06 (0.92-1.22) |  |
| ≥ 69 years | 3599 | 851 | 1.07 (0.99-1.15) |  | 517 | 1.02 (0.93-1.13) |  | 533 | 1.07 (0.97-1.18) |  |
| Race and ethnicity |  |  |  | 0.83 |  |  | 0.50 |  |  | 0.54 |
| Black | 414 | 130 | 1.12 (0.93-1.35) |  | 107 | 1.15 (0.94-1.41) |  | 56 | 0.93 (0.71-1.23) |  |
| Hispanic | 160 | 41 | 1.26 (0.85-1.86) |  | 24 | 1.04 (0.61-1.77) |  | 26 | 1.38 (0.83-2.29) |  |
| White | 5297 | 1096 | 1.06 (0.99-1.13) |  | 639 | 1.02 (0.93-1.11) |  | 686 | 1.08 (0.99-1.18) |  |
| E-alone trial |  |  |  | 0.28 |  |  | 0.33 |  |  | 0.31 |
| E-alone intervention | 1152 | 260 | 1.06 (0.93-1.21) |  | 181 | 1.06 (0.90-1.24) |  | 144 | 0.99 (0.83-1.19) |  |
| E-alone placebo | 1163 | 270 | 1.29 (1.13-1.47) |  | 169 | 1.32 (1.11-1.56) |  | 152 | 1.20 (1.00-1.43) |  |
| E + P trial |  |  |  | 0.83 |  |  | 0.61 |  |  | 0.97 |
| E + P intervention | 1854 | 404 | 1.01 (0.90-1.13) |  | 233 | 0.90 (0.77-1.05) |  | 260 | 1.05 (0.91-1.21) |  |
| E + P placebo | 1900 | 373 | 0.99 (0.88-1.11) |  | 216 | 0.98 (0.84-1.13) |  | 230 | 1.04 (0.90-1.21) |  |
| Menopause type |  |  |  | 0.07 |  |  | 0.02 |  |  | 0.92 |
| Natural | 4753 | 1007 | 1.03 (0.96-1.10) |  | 602 | 0.98 (0.90-1.07) |  | 621 | 1.06 (0.97-1.16) |  |
| Surgical | 451 | 93 | 1.22 (0.99-1.51) |  | 59 | 1.38 (1.06-1.79) |  | 49 | 0.99 (0.71-1.37) |  |
| *APOE ε4* carriage |  |  |  | 0.08 |  |  | 0.68 |  |  | 0.07 |
| 0 *ε4 alleles* | 3858 | 724 | 1.08 (1.00-1.18) |  | 440 | 1.01 (0.91-1.12) |  | 436 | 1.12 (1.01-1.25) |  |
| ≥ 1 *ε4 alleles* | 1306 | 357 | 0.97 (0.86-1.09) |  | 190 | 0.99 (0.84-1.17) |  | 238 | 0.97 (0.84-1.12) |  |
|  | DNAm logCRP | | | | | | | | | |
|  |  | MCI/probable dementia | | | MCI | | | Probable dementia | | |
|  | n | n events | HR (95% CI) | P-int | n events | HR (95% CI) | P-int | n events | HR (95% CI) | P-int |
| Overall | 6069 | 1307 | 1.07 (1.00-1.14) |  | 799 | 1.08 (0.99-1.17) |  | 786 | 1.04 (0.95-1.14) |  |
| Age |  |  |  | 0.05 |  |  | 0.04 |  |  | 0.18 |
| > 69 years | 2470 | 456 | 1.02 (0.90-1.15) |  | 282 | 1.01 (0.87-1.17) |  | 253 | 1.01 (0.86-1.18) |  |
| ≥ 69 years | 3599 | 851 | 1.09 (1.01-1.19) |  | 517 | 1.11 (1.00-1.23) |  | 533 | 1.07 (0.96-1.19) |  |
| Race and ethnicity |  |  |  | 0.14 |  |  | 0.86 |  |  | 0.02 |
| Black | 414 | 130 | 1.04 (0.86-1.25) |  | 107 | 1.10 (0.90-1.35) |  | 56 | 0.81 (0.62-1.07) |  |
| Hispanic | 160 | 41 | 0.90 (0.52-1.57) |  | 24 | 1.18 (0.58-2.4) |  | 26 | 0.53 (0.23-1.22) |  |
| White | 5297 | 1096 | 1.09 (1.01-1.17) |  | 639 | 1.09 (0.99-1.2) |  | 686 | 1.09 (0.99-1.20) |  |
| E-alone trial |  |  |  | 0.98 |  |  | 0.52 |  |  | 0.91 |
| E-alone intervention | 1152 | 260 | 1.04 (0.89-1.21) |  | 181 | 1.11 (0.92-1.33) |  | 144 | 0.98 (0.79-1.21) |  |
| E-alone placebo | 1163 | 270 | 1.14 (0.98-1.32) |  | 169 | 1.15 (0.95-1.39) |  | 152 | 1.08 (0.89-1.32) |  |
| E + P trial |  |  |  | 0.06 |  |  | 0.01 |  |  | 0.23 |
| E + P intervention | 1854 | 404 | 1.02 (0.90-1.15) |  | 233 | 0.97 (0.82-1.15) |  | 260 | 1.04 (0.89-1.22) |  |
| E + P placebo | 1900 | 373 | 1.12 (0.99-1.28) |  | 216 | 1.15 (0.97-1.36) |  | 230 | 1.12 (0.95-1.33) |  |
| Menopause type |  |  |  | 0.81 |  |  | 0.76 |  |  | 0.99 |
| Natural | 4753 | 1007 | 1.06 (0.98-1.15) |  | 602 | 1.06 (0.96-1.17) |  | 621 | 1.04 (0.94-1.15) |  |
| Surgical | 451 | 93 | 1.01 (0.78-1.32) |  | 59 | 1.10 (0.80-1.50) |  | 49 | 0.95 (0.65-1.40) |  |
| *APOE ε4* carriage |  |  |  | 0.96 |  |  | 0.37 |  |  | 0.66 |
| 0 *ε4 alleles* | 3858 | 724 | 1.10 (1.00-1.21) |  | 440 | 1.04 (0.93-1.18) |  | 436 | 1.14 (1.01-1.29) |  |
| ≥ 1 *ε4 alleles* | 1306 | 357 | 1.07 (0.94-1.21) |  | 190 | 1.21 (1.01-1.44) |  | 238 | 0.98 (0.84-1.15) |  |

Abbreviations: HR = hazard ratio, CI = confidence interval.
Models adjusted for age, race/ethnicity, education, hormone therapy trial arm, total energy expenditure from recreational physical activity, BMI, and white blood cell counts (CD8T, CD4T, natural killer cells, B cells, monocytes, and neutrophils).

Models across strata of race and ethnicity, hormone therapy trial arm, menopause type, and *APOE ε4* carriage did not contain the stratification variable.

**Supplemental Methods**

*DNA methylation (DNAm) measurement*

DNAm from whole blood collected at baseline was measured at the University of Minnesota Genomics Center using the Illumina EPIC v2 (Illumina, Inc., San Diego, CA) BeadChip, which measures methylation at ~930,000 cytosine-guanine dinucleotide (CpG) sites.

*DNA Methylation Quality Control*

Samples were assessed for quality using 17 control metrics from Illumina. Samples were removed if they did not meet Illumina’s recommended thresholds for each control metric (N=14 removed). Sex was determined by clustering samples on the average intensity values of CpG sites on the X and Y chromosomes.^1^ Samples that fell outside of the female cluster or had a mismatch between reported and detected sex were excluded (N=8 removed). Samples with mean bisulfite intensity values < 4,000 were excluded (N=1 removed). Detection p-values were calculated using out-of-band (OOB) probes.^2^ CpG measurements were set to missing if detection p > 0.05 or <= 3 detection beads (M=66,532 probes). CpG sites were removed if >5% of samples were missing data. Based on the remaining CpG sites, samples were excluded if >5% of CpG sites had missing methylation values (N=43 removed). In the remaining samples, OOB background correction, RELIC dye bias correction, and RCP probe type bias correction were applied using Enmix.^3^

*Relatedness*

Kinship was determined from genotype data using KING.^4^ For each pair of participants with estimated 3rd degree or closer relatedness, one was excluded, with the preference to retain cases. If case status matched, one of the pairs was removed at random.

*Principal components*

A random subsample of 172,000 CpG sites spanning the genome was extracted. Methylation values were transformed to m-values. Missing CpG data were imputed to the mean. Principal components were calculated in unrelated, non-replicate samples using the big_randomSVD function in the bigstatsr library.^5^ In remaining samples, PC values were estimated via projection.

*Sample concordance*

Concordance between samples was measured using the SNP fingerprinting probes built into the array. Illumina SNP probes were converted to genotype data. These genotypes were assessed for pairwise IBD in PLINK. If a sample had a 100% match with an unexpected sample, the pair was compared against genotype array data. Under the assumption that array genotypes represented the 'truth' dataset, the sample from the pair that did not match the array genotype was excluded.

*Epigenetic clocks*

The online Horvath and Clock Foundation DNA Methylation Age Calculator (<https://dnamage.clockfoundation.org/>) calculated AgeAccelHorvath, AgeAccelHannum, intrinsic epigenetic age acceleration (IEAA), IEAA.Hannum, extrinsic epigenetic age acceleration (EEAA), AgeAccelPheno, AgeAccelGrim2, and the DNAm-based AgeAccelGrim2 componnents: adrenomedullin (DNAm ADM), beta-2 microglobulin (DNAm B2M), cystatin C (DNAm Cystatin C), growth differentiation factor 15 (DNAm GDF-15), leptin (DNAm leptin), smoking pack-years (DNAm PACKYRS), plasminogen activation inhibitor (DNAm PAI-1), tissue inhibitor metalloproteinase 1 (DNAm TIMP-1), log hemoglobin A1C (DNAm logA1C), and log C-reactive protein (DNAm logCRP)^10^. We calculated the PC epigenetic clocks using R code available at (<https://github.com/MorganLevineLab/PC-Clocks>) and subsequently regressed them on chronological age to compute their respective residual values, henceforth referred to as PCHorvath1Resid, PCHannumResid, PCPhenoAgeResid, and PCGrimAgeResid. DunedinPACE was calculated using R code available at (<https://github.com/danbelsky/DunedinPACE>).

AgeAccelHorvath, AgeAccelHannum, IEAA, IEAA.Hannum, and EEAA are referred to as first generation clocks and were trained on chronological age. IEAA was derived with the Horvath epigenetic clock and additionally captures cell-intrinsic aging independent of white blood cell composition. IEAA.Hannum also captures cell-intrinsic aging but was derived from the Hannum epigenetic clock. EEAA captures age-related changes in white blood cell composition as derived from the Hannum clock. AgeAccelPheno estimates phenotypic age, which consists of age and 9 clinical biomarkers (e.g., creatinine), to predict lifespan and healthspan.^13^ AgeAccelGrim2 is a composite biomarker of DNAm-based surrogates of plasma proteins, smoking pack-years, age, and sex, and is a strong predictor of mortality. PC versions of these clocks were developed to reduce technical variation and improve reliability.^14^ DunedinPACE captures the pace of aging across various organ systems (e.g., cardiovascular and metabolic) and was developed using longitudinal data from 19 biomarkers (e.g., blood pressure).^15^ DunedinPACE has been shown to be associated with cognitive function, mortality, and incidence of multiple chronic diseases.^15^
